# Exploring the Gut-Brain Connection in Gastroparesis with Autonomic and Gastric Myoelectric Monitoring

**DOI:** 10.1101/2022.10.21.22281323

**Authors:** Sandya Subramanian, David C. Kunkel, Linda Nguyen, Todd P. Coleman

## Abstract

**Objective:** The goal of this study was to identify autonomic and gastric myoelectric biomarkers from throughout the day that differentiate patients with gastroparesis, diabetics without gastroparesis, and healthy controls, while providing insight into etiology.

**Methods:** We collected 19 24-hour recordings of electrocardiogram (ECG) and electrogastrogram (EGG) data from healthy controls and patients with diabetic or idiopathic gastroparesis. We used physiologically and statistically rigorous models to extract autonomic and gastric myoelectric information from the ECG and EGG data respectively. From these, we constructed quantitative indices which differentiated the different groups and demonstrated their application in automatic classification paradigms and as quantitative summary scores.

**Results:** We identified several differentiators that separate healthy controls from gastroparetic patient groups, specifically around sleep and meals. We also demonstrated the downstream utility of these differentiators in automatic classification and quantitative scoring paradigms. Even with this small pilot dataset, automated classifiers achieved an accuracy of 79% separating autonomic phenotypes and 65% separating gastrointestinal phenotypes. We also achieved 89% accuracy separating controls from gastroparetic patients in general and 90% accuracy separating diabetics with and without gastroparesis. These differentiators also suggested varying etiologies for different phenotypes.

**Conclusion:** The differentiators we identified were able to successfully distinguish between several autonomic and gastrointestinal (GI) phenotypes using data collected while at-home with non-invasive sensors.

**Significance:** Autonomic and gastric myoelectric differentiators, obtained using at-home recording of fully non-invasive signals, can be the first step towards dynamic quantitative markers to track severity, disease progression, and treatment response for combined autonomic and GI phenotypes.

## I. Introduction

**T**he autonomic nervous system is known to be involved in the regulation of normal digestive activity in the stomach through both the sympathetic ‘fight-or-flight’ and parasympathetic ‘rest-and-digest’ arms. Specifically, the parasympathetic arm mainly promotes digestive activity through the vagus nerve, while the sympathetic arm restricts digestive activity via thoracolumbar ganglia [1]. However, this connection between the autonomic nervous system and the stomach can be disrupted or dysfunctional in certain circumstances. Gastroparesis, or delayed emptying of the stomach in the absence of a mechanical obstruction, may be such a circumstance. With a prevalence of 1.5-3%, gastroparesis can occur due to many causes [2]. Two main types of gastropareses are diabetic gastroparesis and idiopathic gastroparesis. Autonomic small-fiber neuropathy is a known part of the progression of diabetes, which may be a contributing factor in diabetic gastroparesis [1]. On the other hand, idiopathic gastroparesis could result in part from autonomic dysfunction, gastric myoelectric dysfunction, or a variety of other contributory causes [2]. Other functional GI diagnoses, such as dumping syndrome (rapid emptying of the stomach) or a physical obstruction preventing digestion, can also involve autonomic dysfunction [1]. Since symptoms can overlap, and symptom resolution does not imply improved gastric emptying [2], specific sub-typing of gastroparesis into etiology is needed for more targeted interventions.

One of the barriers to in-depth characterization of the gut-autonomic connection in gastroparesis is a lack of multimodal ambulatory monitoring of electrophysiological activity at high temporal resolution with respect to the autonomic nervous system and digestive activity. Previous studies attempting to characterize autonomic and digestive activity in gastroparesis were either not ambulatory, not at a high temporal resolution, not high-quality in terms of physiologically and statistically rigorous methods, or not multimodal [1-12]. Several of the aforementioned autonomic studies have focused on heart rate variability, an important marker of autonomic tone [13].

Heart rate variability characterizes sympathetic and parasympathetic activity through beat-to-beat differences in the intervals between heartbeats. It varies throughout the course of the day based on circadian patterns, internal or external stimuli, and ongoing activity, making it difficult to capture systematic differences within a limited window of measurement [14]. It can also vary based on the models used to compute it, especially if such methods are not statistically or physiologically rigorous enough to capture subtle or detailed beat-to-beat differences. Previous studies of gastroparesis involving HRV have often had conflicting or unclear results. For example, both rapid and delayed gastric emptying were found in a cohort of patients with known autonomic dysfunction [15]. Further, the severity of autonomic dysfunction did not predict severity of foregut symptoms or degree of gastric emptying delay [15].

A potential solution to these issues is to combine recent advancements in ambulatory wearable sensing technology with modern physiology-based statistical models for heart rate variability and gastric electrophysiology. A two-pronged multimodal approach combining both autonomic and digestive activity along with 24-hour ambulatory monitoring has the potential to shed light on detailed patterns of gut-autonomic dysfunction difficult to capture using traditional methods in the clinic. Sleep is characterized by unique and specific autonomic activity, making it a rich testbed for autonomic regulation. Similarly, the period of digestion after meals can highlight irregularities in digestive activity. This degree of individualized insight can inform clinical management and personalized treatment plans, including type, timing, and dosage of medication, timing of meals and sleep, and the potential therapeutic value of non-pharmacologic solutions such as vagal nerve stimulation [16].

In this study, we employed such a paradigm in which we collected 19 24-hour ambulatory electrocardiogram (aECG) and ambulatory electrogastrogram (aEGG) recordings from a combination of healthy controls, diabetic gastroparesis patients, idiopathic gastroparesis patients, and diabetic non-gastroparesis patients. Our subject cohort spanned across primarily autonomic (e.g., diabetic) and GI (e.g., gastroparesis, dumping syndrome) phenotypes. In **Methods**, we provide details about our subject cohort and the autonomic and digestive measures we computed using the aECG and aEGG data respectively. We then discuss how we compared patterns of autonomic and gastric activity between the different subject groups throughout the day, specifically focusing on sleep and meal consumption time periods. In **Results**, we identify key differentiators between autonomic and GI phenotypes. We show how these differentiators suggest different etiologies for different autonomic and GI phenotypes, some involving changes to intensity of modulation, and some involving changes to regulatory patterns of modulation. We also demonstrate the potential utility of our approach for other downstream uses, including defining quantitative autonomic and digestive dysfunction “scores” and automated classification of different phenotypes using multimodal information. Finally, in **Discussion**, we explain the implications of our work and suggest future directions.

## II. Methods

### A. Data

Nineteen continuous 24-hour electrocardiogram (ECG), ambulatory electrogastrogram (aEGG), and triaxial accelerometer data were recorded from 13 subjects (ages 22-70, 5 females) under protocol approved by the University of California San Diego Institutional Review Board using an OpenBCI Cyton ambulatory electrophysiological recording system with 8 channels on the upper abdomen [17]. Subjects were also required to manually annotate when they went to sleep and awakened and consumed meals during the 24-hour period. Further details about data collection, experimental setup, and hardware can be found in [17].

### B. Extracting autonomic and gastric motility information

We analyzed the data using two parallel pathways: first the autonomic information in heart rate variability (HRV), and second the gastric myoelectric information in the aEGG. The autonomic and gastric myoelectric information are complementary and non-redundant with respect to physiology. In each case, we focused on quantifying both the intensity of activity and the degree of appropriate regulation of activity. We also used the triaxial accelerometer information along with the manual annotations to infer sleep and wake times.

#### 1) Inferring sleep and wake from actigraphy

Starting from the 24-hour triaxial accelerometer information for each recording as well as the manual annotations, we used the methodology described in [18] to infer accurate sleep and wake times. In most cases, they were only slight deviations from the manually annotated times. However, in some cases, the subjects forgot to manually annotate their sleep and wake times altogether. In those cases, the automatically computed sleep and wake times were obtained.

#### 2) Autonomic information

We captured HRV information from the ECG using a point process statistical model derived from physiology that yields truly instantaneous estimates of both time domain and frequency domain HRV measures [19] and has been validated in a variety of clinical settings, including anesthesia [20]. After extracting the times of R peaks from the ECG using the rpeakdetect package [21] in Python 3.0, we computed several continuous indices using the point process HRV model. Because we were interested in measuring autonomic modulation, we focused mainly on existing frequency domain measures, specifically instantaneous low frequency power (LF), high frequency power (HF), sympathovagal balance (LF/HF), normalized LF (LFnu), and total autonomic modulation (Totpow). High frequency power (0.15-0.40 Hz) represents parasympathetic activity, while low frequency power (0.04-0.15 Hz) represents a combination of both sympathetic and parasympathetic activity [22]. Sympathovagal balance is the ratio of LF to HF; total autonomic modulation is the sum of LF and HF; and normalized LF is the proportion of total autonomic modulation that is LF [22]. We compared the autonomic information during different periods of the day and between groups, specifically focusing on overall intensity of autonomic modulation, regulation of circadian patterns, and sleep-wake modulation. The discussion of how we assessed overall autonomic modulation and sleep vs wake modulation are in the Supplementary Information.

##### Sleep cycles

We hypothesized that during normal sleep, sleep cycles would be mirrored by vagal activity that also cycles in intensity [23]. Sleep cycles have an average period of approximately 90 minutes, with some individual variability [24]. To capture these sleep cycles from HF, we filtered HF below 6e-4 Hz using a multitaper spectrogram [25] with a window length of 2 hours, sliding by 5 minutes. This identifies oscillatory activity with a period longer than 28 minutes. We computed the power in each band during both sleep and wake in decibels (dB) and focused on the band with a period of 87 minutes, the approximate duration of a sleep cycle. We specifically analyzed the average power in that band during sleep (intensity) and the difference in average power in that band between sleep and wake (appropriate regulation).

##### Vagal modulation patterns during sleep

We also wanted to analyze the overall modulatory patterns of vagal activity during sleep, irrespective of intensity or magnitude. To do this, we used a functional data analysis technique called functional principal components analysis (fPCA) [26]. We used the vagal activity during overnight sleep from each recording filtered below 6e-4 Hz and rescaled it to be between 0 and 1 and the same duration (x-axis stretching). Then we performed fPCA and examined the first four functional principal components (fPC’s) as well as the associated scores for each recording of sleep. These scores quantify the degree to which the ‘shape’ of the vagal dynamics during each recording of overnight sleep matches the shape of each fPC. For example, a subject’s fPC1 score quantifies how similar their vagal sleep pattern is to the first fPC. For further details on fPCA, see [26].

#### 3) Gastric myoelectric information

We captured gastric myoelectric information from the aEGG by computing a spectrogram to determine the power in the frequency band around 0.05 Hz, the frequency of the Cajal cells of the human stomach whose action potentials coordinate smooth muscle contractions [27]. Using the same approaches as described in [17], we performed artifact rejection and physiologic signal processing methods to extract this neuromuscular information. Specifically, we recorded the EGG at a sampling rate of 250 Hz, divided the time domain data into consecutive four-minute segments with 75% overlap, and applied a Hamming window to each. We defined the normalized EGG power as the mean power in the 0.04– 0.06 Hz band minus the background noise level in the 0.06– 0.10 Hz band to control for noise variability between recordings and channels. The full details of the data analysis can be found in [17].

Based on the manual annotations of mealtimes and content, we classified meals as either meals or snacks. Then we focused on isolated meals, which were preceded by at least 4 hours of fasting and for which there were at least 3 hours of recorded data after the meal. We had a total of 29 isolated meals across the dataset, with each recording contributing between 1 and 3 isolated meals. For each isolated meal, we computed the area under the normalized EGG power curve (AUC) as described in [28] during the 4-hour post-prandial period and compared it to the AUC of fasting periods. The full details of this computation and comparison are in the Supplementary Information.

##### Gastric electrophysiologic activity dynamics

We additionally used fPCA analysis to assess the modulation patterns of gastric motility during the postprandial period irrespective of intensity or magnitude. To do this, we rescaled the absolute postprandial (4-hour) power around 0.05 Hz after isolated meals between 0 and 1. Since signals must be the same length for fPCA, we rescaled the duration of the one isolated meal for which we had slightly less than 4 hours of postprandial data to be same length as the others. We then performed an fPCA analysis, focusing on the first four fPC’s, and computed the associated scores.

### C. Automatic classification

After comparing each of these measures between groups, we also used them as features in a classifier to determine if we can use them to automatically delineate patients from controls, and specific phenotypes of disease within patient groups from others. For example, we used a subset of the features extracted from the autonomic information covering both intensity and appropriate regulation of activity to classify autonomic phenotypes using each of the 19 recordings, specifically healthy controls vs diabetic vs non-diabetic patients. We used a subset of the features extracted from the gastric motility information (covering both intensity and appropriate regulation of activity) to classify GI phenotypes for each of the 29 isolated meals, specifically healthy controls vs diabetic gastroparesis vs. diabetic non-gastroparesis vs. idiopathic gastroparesis. The features used for classification are indicated with an asterisk in Table 1. In both cases, we used leave-one-recording-out cross-validation and assessed overall performance separating patients from controls as well as the subgroups of patients. For the isolated meals, all the isolated meals from a single recording were used as a held-out test set for each fold. For the fPCA scores, to stay true to leave-one-recording-out cross-validation, in each fold, the fPCA analysis was done using 18 recordings and the held-out recording’s fPC scores (or those of all the isolated meals from that recording) were computed by projection onto the resulting fPCs. Three different classifier types were tested: multinomial regression [29], K-nearest neighbor (KNN) with 1 neighbor [30], and support vector machine (SVM) with a linear kernel (using only the last 5 of the 12 features for SVM) [31]. Deep learning methods were not implemented because of the relatively small size of the dataset. The features used for each classification task are in Table 1.

**TABLE I.** Summary of autonomic and gastric motility features used and their trends in different patient groups

| FEATURES |  |  |
| --- | --- | --- |
| Autonomic features |  | Summary of trend |
| 1 | *Overall LF | Higher for healthy controls than diabetics |
| 2 | *Overall HF | Higher for healthy controls than diabetics |
| 3 | *Overall Totpow | Higher for healthy controls than diabetics |
| 4 | *Difference of median HF Wake - Sleep | Closest to zero for diabetics, most negative for healthy controls |
| 5 | *Difference of median Bal Wake - Sleep | Closest to zero for diabetics, highest for healthy controls |
| 6 | *Difference of median LFnu Wake - Sleep | Closest to zero for diabetics, highest for healthy controls |
| 7 | *Avg vagal power in 87 min band during sleep | Higher for healthy controls than diabetics and idiopathics |
| 8 | *Sleep – wake avg vagal power in 87 min band | Higher for healthy controls than diabetics and idiopathics |
| 9 | *fPC1 score vagal modulation during sleep | High for healthy controls, low for idiopathics |
| 10 | *fPC2 score vagal modulation during sleep | High for diabetics, low for healthy controls and idiopathics |
| 11 | *fPC3 score vagal modulation during sleep | High for idiopathics |
| Gastric motility features |  | Summary of trend |
| 12 | Normalized post-prandial power (4 hrs) | Higher for healthy controls than gastroparetics (without stimulators) |
| 13 | Difference of normalized post-prandial power (4 hrs) from sleep (fasting) | Higher for healthy controls than gastroparetics (without stimulators) |
| 14 | *fPC1 score gastric motility 4 hrs post-prandial | High for idiopathics and non-gastroparetics, low for diabetic gastroparesis |
| 15 | *fPC2 score gastric motility 4 hrs post-prandial | Can vary across all subjects |
| 16 | *fPC3 score gastric motility 4 hrs post-prandial | High for healthy controls, low for gastroparetics and non-gastroparetics |

\*Later used in automatic classification

### D. Defining quantitative dysfunction scores

Using the autonomic and gastric neuromuscular scores, we also defined our own quantitative function scores based on the trends observed in the measures in Table I, focusing on the intensity of and the appropriate regulation of activity. Therefore, we defined four scores: an autonomic intensity score (AIS), an autonomic regulation score (ARS), a gastric neuromuscular intensity score (GNIS), and a gastric neuromuscular regulation score (GNRS). Each score was defined using features relevant to that score. For example, the autonomic intensity score was defined using the levels overall autonomic modulation and the intensity of sleep cycles. In contrast, the autonomic regulation score was defined using the difference in autonomic modulation between sleep and wake, the difference in sleep cycle intensity between sleep and wake, and the qualitative vagal modulatory patterns (fPCA analysis). The autonomic scores were calculated for each 24-hour recording, while the gastric neuromuscular scores were calculated for each isolated meal. The definition of each of the four scores is below:

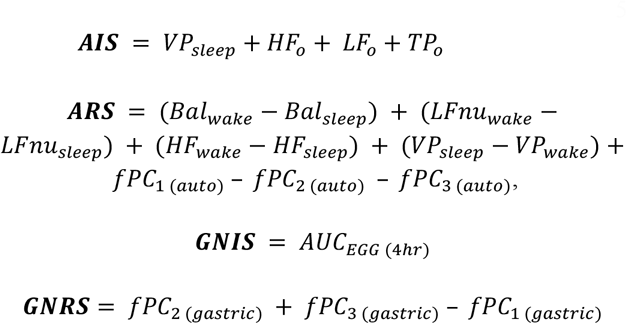

We validated the definition of these scores by showing that each was able to distinguish between relevant autonomic and/or GI phenotypes and provided non-redundant insight into the difference between groups.

## III. Results

An example of processed data from two example recordings are shown in Fig 1. The first is from a healthy control and the second is from a diabetic gastroparesis subject. In each plot, both autonomic and digestive activity markers are shown, including LF/HF, LFnu, and HF for autonomic activity and absolute and normalized power around 0.05 Hz for digestive activity. General periods of wake and sleep as well as meals are marked. Similar plots are shown for all of the recordings in the Supplementary Information (Figs. S3-S21).

**Fig. 1.**
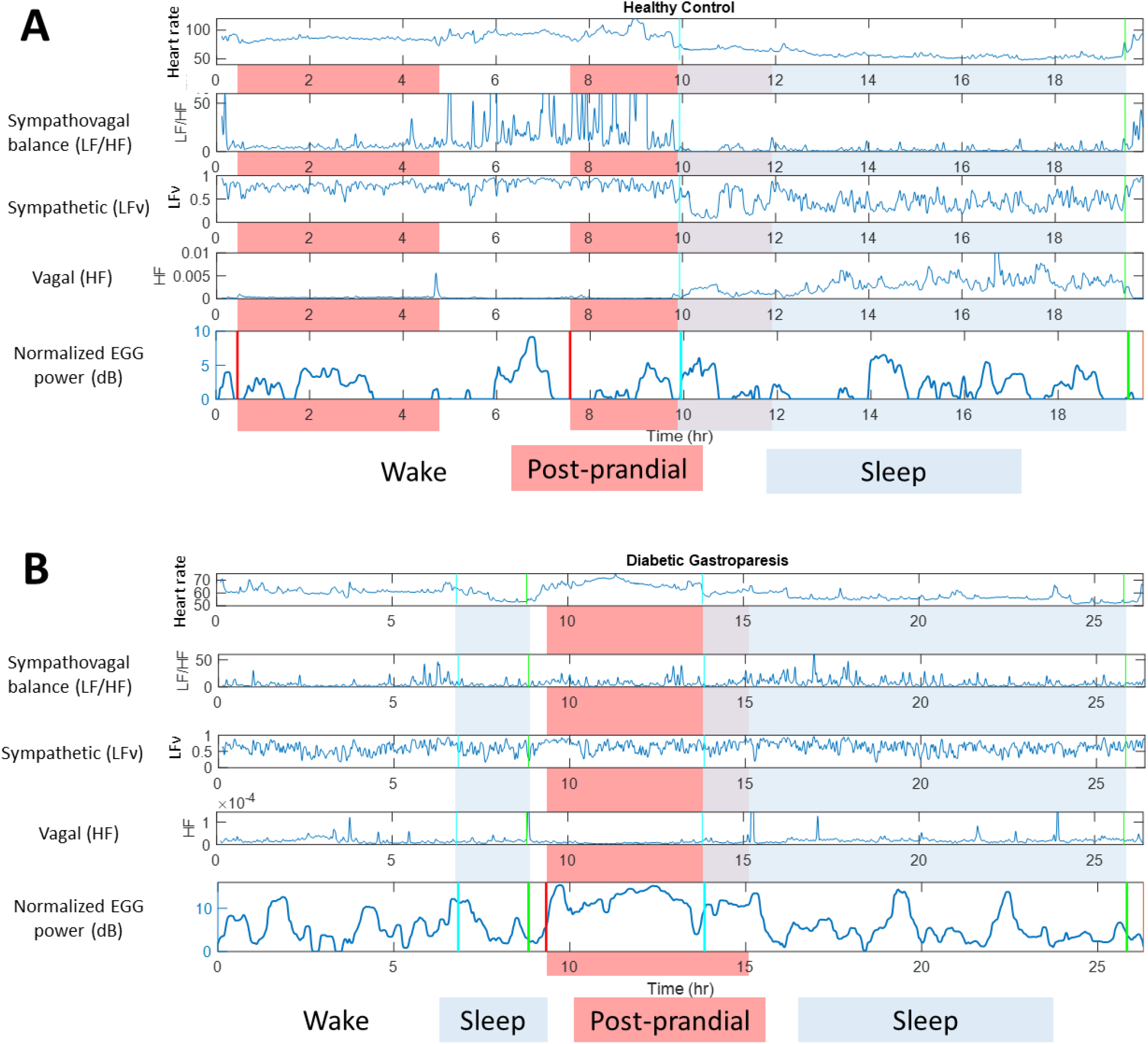
Two examples of autonomic (HRV) and gastric myoelectric information extracted from ECG and EGG data respectively, showing the mean heart rate, standard deviation of heart rate, sympathovagal balance, sympathetic and parasympathetic (vagal) modulation, and normalized EGG power over the course of 24 hours. (A) A healthy control subject, (B) a diabetic gastroparesis patient

### A. Automatically inferring sleep and wake from actigraphy

Overnight sleep and wake times were successfully computed for all 19 24-hour recordings. Even though some subjects may have taken short naps during the day, we focused on overnight sleep to observe circadian autonomic patterns.

### B. Autonomic information

The results of analyzing overall autonomic modulation and wake vs sleep modulation are in the Supplementary Information (Fig S1).

#### 1). Sleep cycles

Fig. 2A shows two examples of spectrograms of vagal activity from two different 24-hour recordings, one from a healthy control and one from a diabetic gastroparesis subject, both on the same color scale for direct comparison. In both cases, the general periods of wake and sleep are marked. The frequency band of interest, with a period of approximately 87 minutes, is the second from the bottom on both spectrograms. In the healthy control spectrogram, there is a clear difference in power in the lower frequency bands between wake and sleep, indicating appropriate regulation of autonomic activity. In contrast, the diabetic subject shows very little systematic difference between wake and sleep. At any time of day, the overall power is much greater for the healthy control than for the diabetic subject, suggesting a difference in general intensity of autonomic activity as well.

**Fig. 2.**
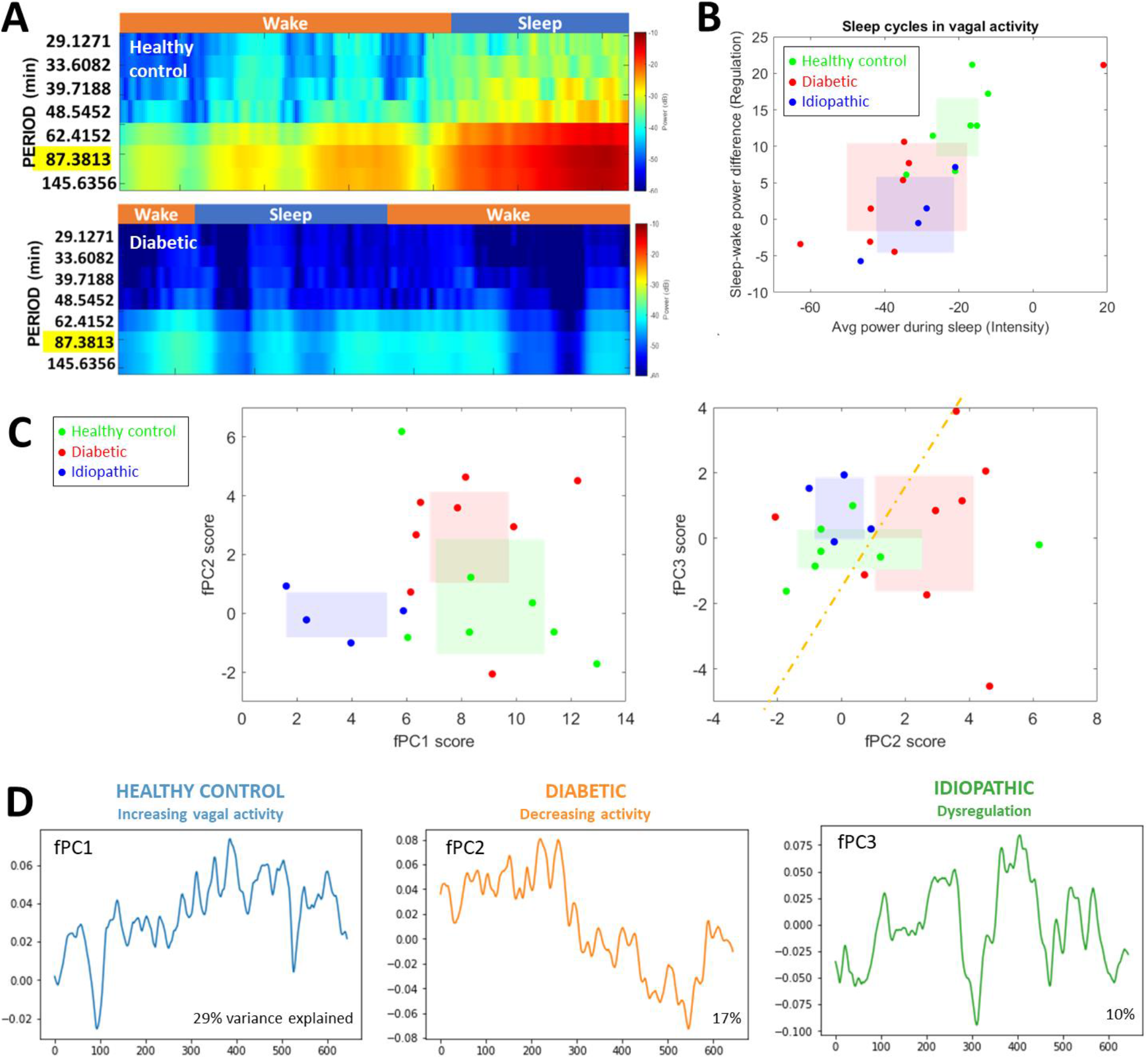
Summary of autonomic activity trends (A) Spectrograms of vagal power in two subjects, a healthy control and a diabetic gastroparesis subject, showing sleep cycle activity (B) Average power of vagal sleep cycles vs difference in vagal sleep cycle power between wake and sleep indicating quality of regulation. (C) Comparison of scores for the first three functional principal components. A dividing line has been added by hand for visual clarity of separation between the diabetics and idiopathics. All colored boxes show the 95% confidence interval of the mean in each dimension for that subgroup of subjects. (D) The vagal modulation pattern captured by each of the first three functional principal components, labeled by which subject subgroup has the highest scores for that fPC and a short description of the modulation pattern

To quantify these differences, Fig. 2B shows the comparison of average power in the 87 min period band during sleep (sleep cycle intensity) and the difference in power in that band between sleep and wake (sleep cycle regulation) for all recordings. The greatest power in that band during sleep and the greatest separation between wake and sleep both occur for healthy controls, who span the top right section the plot. At a group level, both sets of patient groups (diabetics and idiopathic gastroparesis) have decreased power in that band during sleep compared to healthy controls (decreased intensity of activity). Both sets of patient groups also have decreased separation between sleep and wake (impaired regulation). Since this difference is computed by subtracting the power during wake from that during sleep, the healthy controls have a positive difference, indicating that there is much higher power in that band during sleep than during wake, which agrees with the notion of sleep cycles. Interestingly, the two patient groups seem to separate further from each other based on whether they have greater disruption to overall intensity of sleep cycles or sleep-wake regulation. For example, the diabetics seem to be higher on the y-axis but more to the left on the x-axis than the idiopathic gastroparesis subjects. This suggests that they are characterized more by decreased intensity of sleep cycles rather than dysregulation. On the other hand, the idiopathic gastroparesis subjects are lower on the y-axis but slightly to the right of the diabetics on the x-axis, suggested that they are characterized more by dysregulated sleep cycles rather than decreased intensity.

#### 2) Vagal modulation patterns during sleep

Fig. 2C shows the functional principal component (fPC) scores of each of the 24-hour recordings plotted against each other for the first three fPCs. The first plot shows fPC1 score vs fPC2 score. The healthy controls span the lower right section of the plot, while the idiopathic gastroparesis patients span the lower left and the diabetics the top half of the plot. This suggests that the diabetics have the highest fPC2 scores, the healthy controls have high fPC1 scores and low fPC2 scores, and the idiopathic gastroparesis subjects have low fPC1 and low PC2 scores. The second plot shows fPC2 score vs fPC3 score. This plot separates the diabetics from the idiopathic gastroparesis subjects. Here, the diabetics span the lower right half of the plot, while the idiopathic gastroparesis subjects span the upper left. Once again, this verifies that the diabetics have the highest fPC2 scores, but it also shows that the idiopathic gastroparesis subjects have the highest fPC3 scores. Based on these plots, each of the first three fPC’s characterize one of the three subgroups: fPC1 scores are highest for healthy controls, fPC2 scores are highest for diabetics, and fPC3 scores are highest for idiopathic gastroparesis subjects.

Fig. 2D shows the patterns of vagal activity during sleep represented by the first three functional principal components. The first fPC, which explains 29% of the variance, shows increasing vagal activity throughout the duration of sleep. This fPC has the highest scores for healthy controls. The second fPC, which explains 17% of the variance, shows decreasing vagal activity through the course of sleep, and specifically a decreased intensity of activity for most of the duration of sleep. This fPC has the highest scores for the diabetics. The third fPC, which explains 10% of the variance, shows a variable pattern of vagal activity with several peaks and troughs. While the intensity of activity overall is not markedly reduced, the pattern of activity is highly variable. This fPC has the highest scores for the idiopathic gastroparesis subjects. This difference in vagal dynamics over the course of sleep between the different subgroups, specifically in terms of both intensity and regulatory pattern, may relate to underlying dysfunction.

### C) Gastric neuromuscular information

The results of analyzing postprandial EGG activity and compared EGG activity between postprandial and fasting periods is in the Supplementary Information (Fig. S2). Fig. 3A shows the functional principal component (fPC) scores of each of the isolated meals plotted against each other for the first three fPCs. The first plot shows fPC1 score vs fPC2 score. The healthy controls and diabetic gastroparesis subjects span the top left section of the plot, while the idiopathic gastroparesis and diabetic non-gastroparesis meals stay very close to the rightmost and bottommost sections of the plot. This suggests that the idiopathic gastroparesis and diabetic non-gastroparesis meals have the highest fPC1 scores, and at a group level, lower fPC2 scores than the healthy controls and diabetic gastroparesis subjects.

**Fig. 3.**
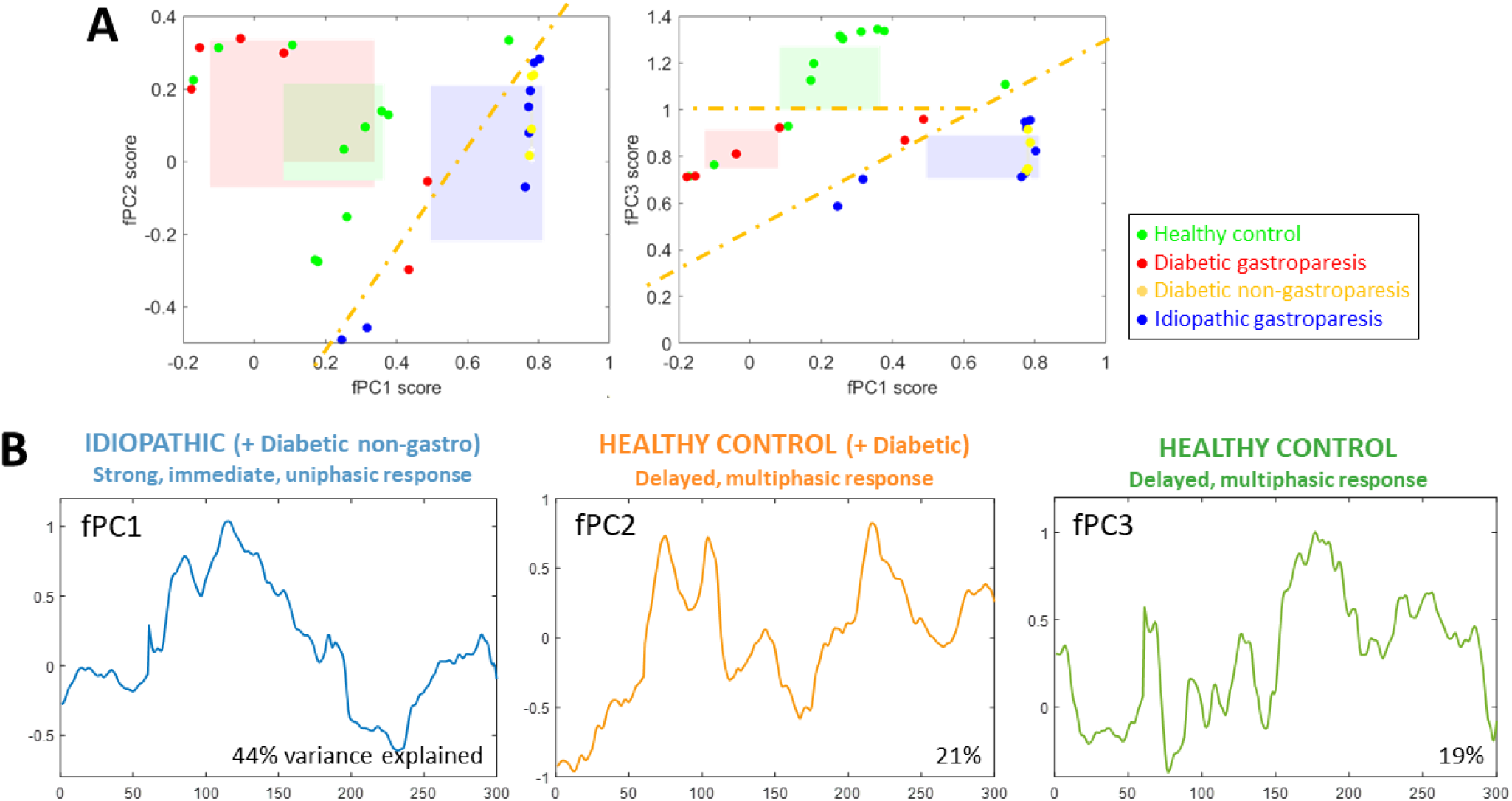
Summary of gastric myoelectric activity patterns. (A) Comparison of scores for the first three functional principal components. All colored boxes show the 95% confidence interval of the mean in each dimension for that subgroup of subjects. (B) The EGG modulation pattern captured by each of the first three functional principal components, labeled by which subject subgroup has the highest scores for that fPC and a short description of the modulation pattern

**Fig. 4.**
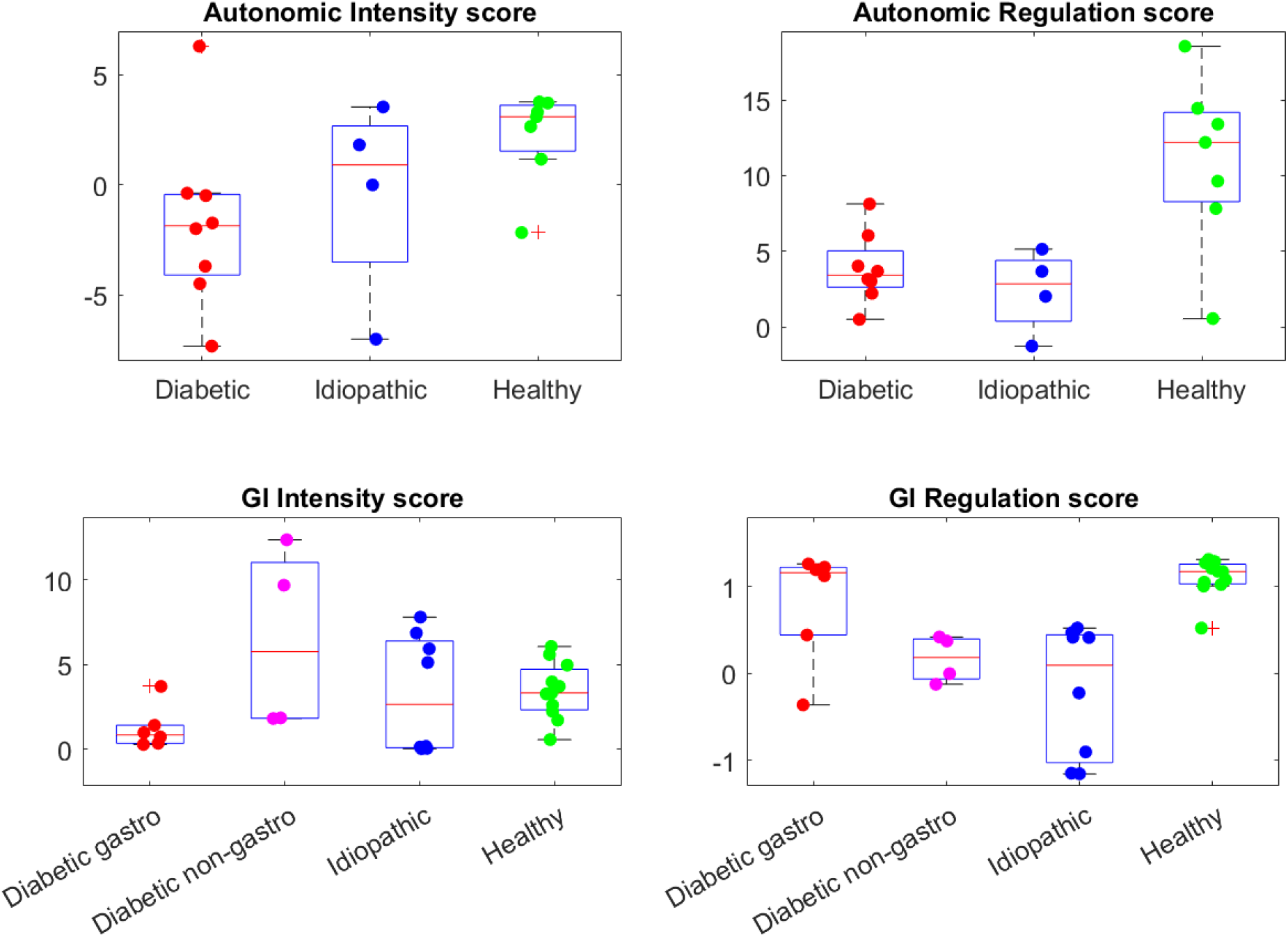
Boxplots of the different diagnostic scores we have developed in this work, separated by subject subgroup.

The second plot shows fPC1 score vs fPC3 score. Here the healthy controls span the top half of the plot, while the diabetic gastroparesis subjects span the lower left side, and the idiopathic gastroparesis subjects and diabetic non-gastroparesis subjects span the lower right side. Once again, this verifies that the idiopathic gastroparesis and diabetic non-gastroparesis subjects have the highest fPC1 scores, but also shows that fPC3 scores separate healthy controls from the patients overall. Healthy controls have the highest fPC3 scores, while all the groups of patients have moderate to low fPC3 scores.

Fig. 3B shows the patterns of postprandial digestive activity represented by the first three functional principal components. The first fPC, which explains 44% of the variance, shows strong digestive activity in the first 2 hours after a meal, with a unimodal peak at around 1 hour postprandially. This fPC has the highest scores for idiopathic gastroparesis and diabetic non-gastroparesis patients. The second fPC, which explains 21% of the variance, shows bimodal digestive activity that peaks first within the first hour after the meal and then again even higher approximately 3 hours after the meal with a trough in between. This fPC has the highest scores at a group level for healthy control and diabetic gastroparesis patients, though some idiopathic gastroparesis subjects also had moderate scores for this fPC. The third fPC, which explains 19% of the variance, shows a multiphase pattern activity with several peaks, first immediately after the meal and then the highest peak around 2 hours after staying increased even 3 hours after the meal. This fPC has the highest scores for healthy controls. Both fPC2 and fPC3, which are most characteristic of healthy controls, are notable for complex multiphase patterns of digestive activity with multiple peaks and the strongest peaks occurring several hours after the meal rather than immediately, which could relate to the phase of the food (solid vs liquid) [32]. In contrast, fPC1, which strongly represents the idiopathic and diabetic non-gastroparesis patients, represents strong digestive activity immediately after the meal without later peaks. This difference in timing and phases of digestive activity may relate to underlying regulatory dysfunction.

### D. Automatic classification

Table 1 summarizes all the features defined across autonomic and gastric motility activity, with the key observations about each.

#### 1) Autonomic-only classification

Using the 11 autonomic features in Table I, we tested multinomial regression and KNN models to differentiate between three autonomic phenotypes, specifically healthy control, diabetic, and non-diabetic (idiopathic) gastroparesis.

**TABLE II.** Summary of autonomic and gastric motility features used and their trends in different patient groups

| Variable | Definition |
| --- | --- |
| $HF_o$ | overall HF |
| $LF_o$ | overall LF |
| $TP_o$ | overall total autonomic modulation |
| $VP_{sleep}$ | average vagal power in the sleep cycle band during sleep |
| $(Bal_{wake} - Bal_{sleep})$ | difference in median sympathovagal balance between sleep and wake |
| $(LFnu_{wake} - LFnu_{sleep})$ | difference in median LFnu between sleep and wake |
| $(HF_{wake} - HF_{sleep})$ | difference in median HF between sleep and wake |
| $(VP_{sleep} - VP_{wake})$ | difference in the vagal power in the sleep cycle band between sleep and wake |
| $fPC_1(auto)$ | autonomic fPC1 score |
| $fPC_2(auto)$ | autonomic fPC2 score |
| $fPC_3(auto)$ | autonomic fPC3 score |
| $AUC_{EGG(4hr)}$ | area under the curve of the normalized EGG power for 4 hours postprandially |
| $fPC_1(gastric)$ | gastric fPC1 score |
| $fPC_2(gastric)$ | gastric fPC2 score |
| $fPC_3(gastric)$ | gastric fPC3 score |
| $AIS$ | Autonomic intensity score |
| $ARS$ | Autonomic regulation score |
| $GNIS$ | Gastric neuromuscular intensity score |
| $GNRS$ | Gastric neuromuscular regulation score |

We also trained an SVM model using a subset of only 5 of the 12 features for the same task. With leave-one-recording-out cross-validation, the best performance achieved was 79% accuracy or 15/19 using either multinomial regression or SVM (both achieved the same accuracy). K-nearest neighbor did slightly worse, correctly classifying 14/19 of the recordings. As summarized in Table III, by group, the accuracy was 86% (6/7) for healthy controls, 75% (6/8) for diabetic gastroparesis, and 75% (3/4) for idiopathic gastroparesis. When only examining the accuracy in terms of distinguishing healthy controls from patients, the best classifier achieved 89% accuracy, or 17/19 correct classifications (Table IV).

**TABLE III.**
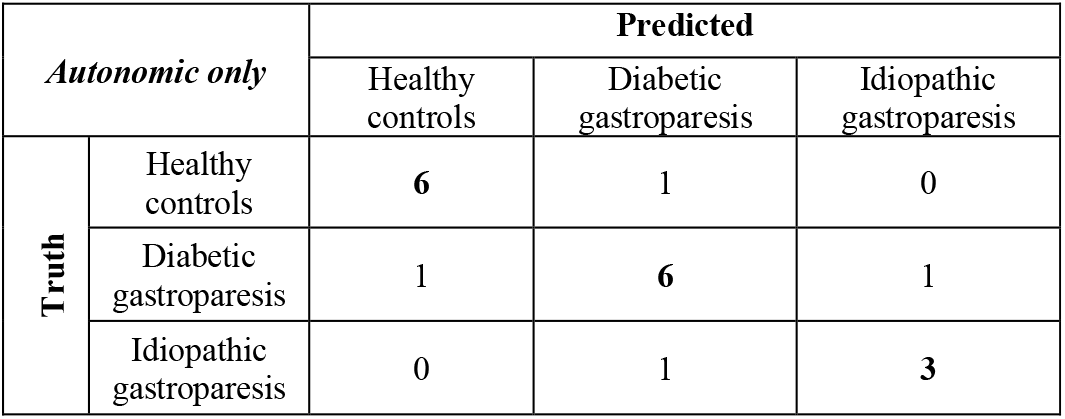
Summary of Automatic Classification Results: Autonomic Only

**TABLE IV.** Summary of Automatic Classification Results

| Classification | Multinomial regression (best performance) |
| --- | --- |
| *Autonomic only* |  |
| Healthy controls v. patients | 17/19 (0.89) |
| *EGG only* |  |
| Healthy controls v. patients | 22/29 (0.76) |
| Diabetic gastroparesis v. diabetic non-gastroparesis | 9/10 (0.90) |

#### 2) EGG-only classification

Using the 3 EGG features in Table 1, we trained and tested a multinomial regression model to differentiate between four GI phenotypes, specifically healthy control, diabetic gastroparesis, diabetic non-gastroparesis, and idiopathic gastroparesis. Gastric stimulators were not treated as a separate phenotype because of very few subjects. Both SVM and KNN performed equally or more poorly compared to multinomial regression. With leave-one-recording-out cross-validation, the best performance achieved was 66% accuracy or 19/29 correct classifications. As summarized in Table V, by group, the accuracy was 73% (8/11) for healthy controls, 33% (2/6) for diabetic gastroparesis, 100% (4/4) for diabetic non-gastroparesis, and 63% (5/8) for idiopathic gastroparesis. Specifically, when assessing accuracy for distinguishing patients from controls, the best classifier achieved 76% accuracy, or 22/29 correct classifications (Table IV). However, the classifier achieved high performance when differentiating between GI-specific phenotypes, such diabetic gastroparesis vs diabetic non-gastroparesis (9/10 or 90% accuracy).

**TABLE V.** Summary of Automatic Classification Results: EGG Only

| *EGG only* |  | Predicted |  |  |  |
| --- | --- | --- | --- | --- | --- |
|  |  | Healthy controls | Diabetic gastroparesis | Diabetic non-gastroparesis | Idiopathic gastroparesis |
| Truth | Healthy controls | 8 | 3 | 0 | 0 |
|  | Diabetic gastroparesis | 4 | 2 | 0 | 0 |
|  | Diabetic non-gastroparesis | 0 | 0 | 4 | 0 |
|  | Idiopathic gastroparesis | 0 | 1 | 2 | 5 |

#### 3) Combined classification

Combining both autonomic and gastric myoelectric features did not improve classification performance further at this stage, likely due to the limited number of recordings compared to the number of phenotypes.

### E. Defining quantitative dysfunction scores

Figure 5 shows the 4 quantitative functional scores that we defined, with 2 focused on autonomic function and 2 focused on gastric motility, for each of the relevant phenotypes (either autonomic or GI). The autonomic intensity scores and autonomic regulation scores are shown for the three autonomic phenotypes (healthy, diabetic, and idiopathic), while the gastric motility intensity score and gastric motility regulation scores are shown for the four GI phenotypes (healthy, diabetic gastroparesis, diabetic non-gastroparesis, idiopathic gastroparesis).

Figure 5 shows that the autonomic scores can distinguish between the autonomic phenotypes, while the gastric motility scores are able to distinguish between specific GI phenotypes. For example, Fig. 5A shows that the autonomic intensity score is highest for the healthy controls as a group and lowest for the diabetics, with the idiopathic subgroup in between. Fig. 5B shows that the autonomic regulation score is highest for the healthy controls, lowest for the idiopathic patients, and only slightly higher for the diabetic patients. These quantify our observations that diabetics tend to have decreased intensity of activity and dysregulation, while idiopathic gastroparesis patients tend to have more dysregulation than decreased intensity. Similarly, the gastric motility intensity score is higher for all three groups except for the diabetic gastroparesis group. The gastric motility regulation score is higher for the healthy controls and diabetic gastroparesis patients than for the diabetic non-gastroparesis and idiopathic gastroparesis patients. Taken together, the two gastric neuromuscular scores can separate diabetic gastroparesis from diabetic non-gastroparesis patients, diabetic gastroparesis from idiopathic gastroparesis, and all the patients from controls. Combining the autonomic and gastric motility scores can give even more nuanced information to separate intersecting autonomic and GI phenotypes. These scores allow us to quantify the group-level observations about the intensity and dysregulation of autonomic and GI activity of the different subgroups, and they can be computed for an individual.

## IV. Discussion

In this paper, we collected 19 24-hour ambulatory ECG and EGG recordings from healthy controls, diabetic gastroparesis patients, idiopathic gastroparesis patients, and diabetic non-gastroparesis patients, and defined autonomic and digestive measures that differentiated subject groups. These autonomic and digestive measures focused on sleep-wake differences and post-prandial periods and successfully distinguished between patients and controls, as well as subgroups of patients with specific autonomic and GI phenotypes. The performance of classification based upon autonomic features was particularly noteworthy. These findings also suggest capabilities to uncover different etiologies with different autonomic and GI phenotypes, such as decreases in intensity of activity or dysregulated patterns of activity. Using these measures, we trained classifiers to separate different phenotypes and defined intensity and regulation ‘scores’ to quantify dysfunction for individual recordings.

There are three main takeaways from this work. First, using statistically rigorous methods, we were able to identify key differentiators (both autonomic and gastric) that are non-redundant with respect to each other and that can distinguish different autonomic and GI phenotypes. These differentiators were primarily associated with the overall intensity of autonomic and gastric activity, circadian patterns of sleep-wake regulation of autonomic activity, and pre-and post-prandial regulation of gastric activity. To compute these differentiators, we employed both time and frequency-based methods and used physiologically validated statistical models for the relevant signals. The autonomic and gastric information are complementary and help delineate autonomic and GI phenotypes respectively. In future work, we will experiment with different approaches to best combine them. Second, we demonstrated proof-of-concept potential downstream use cases for these identified differentiators, including to train automated classifiers and define quantitative scores. Eventually, they can help define new objective quantitative markers for measuring severity and monitoring progression or improvement in response to treatment in gastroparesis and other functional GI disorders. Similar findings were recently published showing that sleep vs wake heart rate variability can be used to predict the apnea hypopnea index in obstructive sleep apnea [33].

Finally, these differentiating markers suggest different primary etiologies for different diagnoses, some more associated with decreased intensity of activity and others more strongly associated with dysregulated activity. For example, diabetic gastroparesis is more strongly associated with decreased intensity of autonomic and gastric activity than idiopathic gastroparesis, while idiopathic gastroparesis has a stronger pattern of dysregulation compared to any changes in intensity of activity. Gastroparesis also has a distinctly different signature than non-gastroparesis diagnoses. This agrees with previous studies showing decreased autonomic modulation and dysregulated motility in functional gastrointestinal disorders (both rapid and delayed gastric emptying and functional dyspepsia) and diabetes, including decreased vagal tone specifically [1-6, 34-38], but our results demonstrate this on an individual basis. The level of nuance and detail obtained in this approach is only possible because of the availability of high-resolution wearable sensors for ambulatory monitoring and the physiology-based statistical models used for computing the initial heart rate variability and gastric neuromuscular indices.

There are limitations to this study. The relatively small number of recordings limited the degree of precision and nuance we could achieve in automatic classification. For example, we could not capture any performance gain from combining features, which would likely be possible in a larger dataset. The classification performed, while a proof-of-concept, is suboptimal because each recording was treated as though it was independent, even though there are three subjects who contributed more than one recording. In our study, we could not separate certain phenotypes, such as the presence of a gastric stimulator, because there were too few recordings. Finally, relying solely on manual annotation for the timing of meals is not ideal since human error and imprecise memory make it challenging to compute the precise pre and post prandial periods accurately. In our future work, we will validate this analysis in a larger cohort of more subjects, in which a richer set of phenotypes and the optimal effects of combining modalities can be studied.

## Data Availability

All data produced in the present study are available upon reasonable request to the authors

## REFERENCES

[1] L. Nguyen et al. (2020). “Autonomic function in gastroparesis and chronic unexplained nausea and vomiting: Relationship with etiology, gastric emptying, and symptom severity”, Neurogastroenterology and Motility 32: e13810.

[2] A. A. Gharibans, T. P Coleman, H. Mousa, and D. C. Kunkel (2019). “Spatial Patterns From High-Resolution Electrogastrography Correlate with Severity of Symptoms in Patients with Functional Dyspepsia and Gastroparesis”, CGH 17: 2668–2677.

[3] T. L. Abell, S. Cardoso, J. Schwartzbaum, B. Familoni, R. Wilson, D. Massie (1994). “Diabetic gastroparesis is associated with an abnormality in sympathetic innervation,” European Journal of Gastroenterology & Hepatology 6(3):241–248.

[4] M. K. Mohammad, D. J. Pepper, A. Kedar, F. Bhaijee, B. Familoni, H. Rashed, T. Cutts, T. L. Abell (2016). “Measures of Autonomic Dysfunction in Diabetic and Idiopathic Gastroparesis,” Gastroenterology Res. 9(4-5):65–69. doi: 10.14740/gr713w. Epub 2016 Sep 20. PMID: 27785328; PMCID: PMC5040547.

[5] M. Heitkemper, M. Jarrett, K. C. Cain, R. Burr, R. L. Levy, A. Feld, V. Hertig (2001). “Autonomic nervous system function in women with irritable bowel syndrome,” Dig Dis Sci. 46(6):1276–84. doi: 10.1023/a:1010671514618. PMID: 11414305.

[6] H. Rashed, T. L. Abell, B. O. Familoni, S. Cardoso (1999). “Autonomic function in cyclic vomiting syndrome and classic migraine,” Dig Dis Sci. 44(8 Suppl):74S-78S. PMID: 10490043.

[7] E. R. Muth, K. L. Koch, R. M. Stern (2000). “Significance of autonomic nervous system activity in functional dyspepsia,” Dig Dis Sci. 45(5):854–63. doi: 10.1023/a:1005500403066. PMID: 10795745.

[8] A. Stocker, T. L. Abell, H. Rashed, A. Kedar, B. Boatright, J. Chen (2016). “Autonomic Evaluation of Patients with Gastroparesis and Neurostimulation: Comparisons of Direct/Systemic and Indirect/Cardiac Measures,” Gastroenterology Res. 9(1):10–16. doi: 10.14740/gr667w. Epub 2016 Mar 8. PMID: 27785318; PMCID: PMC5051107.

[9] G. N. Verne, C. Soldevia-Pico, M. E. Robinson, K. M. Spicer, A. Reuben (2004). “Autonomic dysfunction and gastroparesis in cirrhosis,” J Clin Gastroenterol. 38(1):72–6. doi: 10.1097/00004836-200401000-00015. PMID: 14679331.

[10] K. Keresztes, I. Istenes, A. Folhoffer, P. L. Lakatos, A. Horvath, T. Csak, P. Varga, P. Kempler, F. Szalay (2004). “Autonomic and sensory nerve dysfunction in primary biliary cirrhosis,” World J Gastroenterol. 10(20):3039–43. doi: 10.3748/wjg.v10.i20.3039. PMID: 15378789; PMCID: PMC4576268.

[11] A. A. Gharibans, S. Calder, C. Varghese, S. Waite, G. Schamberg, et al. (2022). “Gastric dysfunction in patients with chronic nausea and vomiting syndromes defined by a noninvasive gastric mapping device,” Science Translational Medicine 14(663). doi: 10.1126/scitranslmed.abq3544.

[12] A. S. Agrusa, D. C. Kunkel, T. Coleman (2022). “Robust Regression and Optimal Transport Methods to Predict Gastrointestinal Disease Etiology from High Resolution EGG and Symptom Severity”, IEEE Trans Biomed Eng. doi: 10.1109/TBME.2022.3167338

[13] “Heart rate variability. Standards of measurement, physiological interpretation, and clinical use. Task Force of the European Society of Cardiology and the North American Society of Pacing and Electrophysiology,” Eur Heart J. 1996;17(3):354–381. doi: 10.1093/oxfordjournals.eurheartj.a014868.

[14] F. Shaffer, J. P. Ginsberg (2017). “An Overview of Heart Rate Variability Metrics and Norms,” Front Public Health. 5:258. doi: 10.3389/fpubh.2017.00258. PMID: 29034226; PMCID: PMC5624990.

[15] A. Lawal, A. Barboi, A. Krasnow, R. Hellman, S. Jaradeh, B. T. Massey (2007). “Rapid gastric emptying is more common than gastroparesis in patients with autonomic dysfunction,” Am J Gastroenterol. 102(3):618–23. doi: 10.1111/j.1572-0241.2006.00946.x. PMID: 17100966.

[16] A. Gottfried-Blackmore, E. P. Adler, N. Fernandez-Becker, J. Clarke, A. Habtezion, L. Nguyen (2020). “Open-label pilot study: Non-invasive vagal nerve stimulation improves symptoms and gastric emptying in patients with idiopathic gastroparesis,” Neurogastroenterol Motil. 32(4): e13769. doi: 10.1111/nmo.13769.

[17] A. A. Gharibans, B. L. Smarr, D. C. Kunkel, L. J. Kriegsfeld, H. M. Mousa, and T. P. Coleman (2018). “Artifact rejection methodology enables continuous, noninvasive measurement of gastric myoelectric activity in ambulatory subjects,” Scientific reports 8(1): 1–12.

[18] (In press) S. Subramanian, T. P. Coleman (2022). “Automated classification of sleep and wake from single day triaxial accelerometer data,” Proc. 44th IEEE International Conf on Eng in Bio and Med (EMBC).

[19] R. Barbieri, E. C. Matten, A. A. Alabi, E. N. Brown (2005). “A point-process model of human heartbeat intervals: new definitions of heart rate and heart rate variability,” Am J Physiol Heart Circ Physiol. 288(1):H424–35. doi: 10.1152/ajpheart.00482.2003. PMID: 15374824.

[20] S. Subramanian, P. L. Purdon, R. Barbieri, E. N. Brown (2021). “Quantitative Assessment of the Relationship between Behavioral and Autonomic Dynamics during Propofol-Induced Unconsciousness,” PLOS ONE 16(8): e0254053. https://doi.org/10.1371/journal.pone.0254053

[21] https://github.com/tru-hy/rpeakdetect

[22] “Heart Rate Variability: Standards of Measurement, Physiological Interpretation, and Clinical Use.” Circulation 93.5 (1996): 1043–1065.

[23] R. Cabiddu, S. Cerutti, G. Viardot, S. Werner, A. M. Bianchi (2012). “Modulation of the Sympatho-Vagal Balance during Sleep: Frequency Domain Study of Heart Rate Variability and Respiration,” Front Physiol. 3:45. doi: 10.3389/fphys.2012.00045. PMID: 22416233; PMCID: PMC3299415.

[24] A. K. Patel, V. Reddy, J. F. Araujo (2022). Physiology, Sleep Stages. [Updated 2022 Apr 28]. In: StatPearls [Internet]. Treasure Island (FL): StatPearls Publishing; Available from: https://www.ncbi.nlm.nih.gov/books/NBK526132/

[25] M. J. Prerau, M. T. Bianchi, R. E. Brown, J. M. Ellenbogen, P. L. Purdon (2017). “Sleep Neurophysiological Dynamics Through the Lens of Multitaper Spectral Analysis,” Physiology 32(1):60-92. PubMed PMID: 27927806.

[26] J. O. Ramsay, B. W. Silverman (2006). Functional data analysis, 2nd edn. Springer Series in Statistics. New York, NY: Springer

[27] K. Sanders, S. Koh, S. Ro et al (2012). “Regulation of gastrointestinal motility—insights from smooth muscle biology,” Nat Rev Gastroenterol Hepatol 9:633–645. https://doi-org.stanford.idm.oclc.org/10.1038/nrgastro.2012.168

[28] Kurniawan, J. F., Tjhia, B., Wu, V. M., Shin, A., Sit, N. L., Pham, T., … & Coleman, T. P. (2021). An Adhesive-Integrated Stretchable Silver-Silver Chloride Electrode Array for Unobtrusive Monitoring of Gastric Neuromuscular Activity. Advanced Materials Technologies, 6(5), 2001229.

[29] A. Sharma (2020). “ML from Scratch-Multinomial Logistic Regression,” Towards Data Science, https://towardsdatascience.com/ml-from-scratch-multinomial-logistic-regression-6dda9cbacf9d

[30] O. Harrison (2018). “Machine Learning Basics with the K-Nearest Neighbors Algorithm,” Towards Data Science, https://towardsdatascience.com/machine-learning-basics-with-the-k-nearest-neighbors-algorithm-6a6e71d01761

[31] R. Gandhi (2018). “Support Vector Machine – Introduction to Machine Learning Algorithms,” Towards Data Science, https://towardsdatascience.com/support-vector-machine-introduction-to-machine-learning-algorithms-934a444fca47

[32] M. Camilleri, J. R. Malagelada, M. L. Brown, G. Becker, and A. R. Zinsmeister (1985). “Relation between antral motility and gastric emptying of solids and liquids in humans”, American Journal of Physiology-Gastrointestinal and Liver Physiology 249(5): G580–G585.

[33] E. C. Nam, K. J. Chun, J. Y. Won, J. W. Kim, W. H. Lee (2022). “The differences between daytime and nighttime heart rate variability may usefully predict the apnea-hypopnea index in patients with obstructive sleep apnea,” J Clin Sleep Med 18(6):1557–1563. doi: 10.5664/jcsm.9912. PMID: 35088710; PMCID: PMC9163618.

[34] R. R. Arora, R. J. Bulgarelli, S. Ghosh-Dastidar, J. Colombo (2008). “Autonomic mechanisms and therapeutic implications of postural diabetic cardiovascular abnormalities,” J Diabetes Sci Technol. 2(4):645–57. doi: 10.1177/193229680800200416. PMID: 19885241; PMCID: PMC2769753.

[35] C. Brock, N. Jessen, B. Brock, P. E. Jakobsen, T. K. Hansen, J. M. Rantanen, S. Riahi, Y. K. Dimitrova, A. Dons-Jensen, Q. Aziz, A. M. Drewes, A. D. Farmer (2017). “Cardiac vagal tone, a non-invasive measure of parasympathetic tone, is a clinically relevant tool in Type 1 diabetes mellitus,” Diabet Med. 34(10):1428–1434. doi: 10.1111/dme.13421. Epub 2017 Aug 17. PMID: 28703868.

[36] D. O’Malley (2016). “Neuroimmune Cross Talk in the Gut. Neuroendocrine and neuroimmune pathways contribute to the pathophysiology of irritable bowel syndrome,” Am J Physiol Gastrointest Liver Physiol 311: G934–G941. doi:10.1152/ajpgi.00272.2016.

[37] B. Salvioli, G. Pellegatta, M. Malacarne, F. Pace, M. Malesci, M. Pagani, D. Lucini (2015). “Autonomic nervous system dysregulation in irritable bowel syndrome,” Neurogastroenterol Motil 27:423–430. doi: 10.1111/nmo.12512

[38] A. E. Bharucha, M. Camilleri, P. A. Low, A. R. Zinsmeister (1993). “Autonomic dysfunction in gastrointestinal motility disorders,” Gut 34(3):397–401. doi: 10.1136/gut.34.3.397. PMID: 8472990; PMCID: PMC1374149.

