## Supplementary Material for "Exploring the Gut-Brain Connection in Gastroparesis with Autonomic and Gastric Myoelectric Monitoring"

**Additional Methods**

Overall autonomic modulation: First we compared overall autonomic modulation intensity throughout the full duration of each recording. To do this, we computed the median of each of the five HRV measures (LF, HF, LF/HF, LFnu, Totpow) for each full recording. We also computed a 95% confidence interval for each median using an empirical bootstrap procedure, which is described in the section below.

Wake vs sleep autonomic modulation: Then, we compared the regulation of autonomic modulation during sleep to that during wake for each recording. Generally, it is known that sleep is dominated by parasympathetic activity compared to wake in healthy subjects. To do this comparison, we computed the median of each of the HRV measures during periods of sleep and periods of wake for each recording. We once again computed confidence intervals for each median empirically. We also computed the difference between the medians of each measure during wake and sleep for each recording and computed empirical confidence intervals for the difference.

We sought to investigate autonomic activity during sleep further, since it is highest in intensity during sleep compared to wake in healthy people. To do this, we focused specifically on parasympathetic or vagal activity, computed by HF.

Empirical bootstrap: We computed empirical confidence intervals for the median values of the HRV and gastric myoelectric measures using the following procedure. For each recording, we sampled with replacement from the full set of values of that index during the recording to create 1,000,000 datasets of the same length as the recording. We computed the median of each of those artificial datasets to construct and empirical distribution for the median. We took the 2.5$^{th}$ and 97.5$^{th}$ percentile values of the empirical distribution as the 95% confidence interval.

Postprandial EGG activity: We computed the area under the curve of the normalized postprandial EGG power during the 4 hours after each isolated meal across all the recordings.

EGG activity in postprandial vs. fasting periods: We compared the area under the curve of normalized postprandial power [1] for each isolated meal to the average power around 0.05 Hz during the last 3 hours of overnight sleep, as a marker of fasting activity. Since not all subjects had well-separated meals, and since annotation of mealtimes was performed manually, using the time immediately before an annotated meal as fasting can be inaccurate. We computed the difference in average power between the postprandial period and the surrogate fasting period for all isolated meals.

**Additional Results**

*Overall autonomic modulation*

Fig. S1A shows the median HF and LF overall over a 24-hour period of each recording. The healthy controls have higher overall intensities of autonomic modulation across all three metrics compared to the diabetics, which agrees with known physiology of diabetes [2]. It is interesting to note that the only healthy control with similar intensity of autonomic modulation as the diabetics is in their 80s, which suggests that like diabetes, aging may affect the autonomic nervous system to reduce intensity of modulation. The idiopathic gastroparesis patients have varying overall intensities, some higher and some lower.

*Wake vs sleep*

Fig. S1B shows the difference in various metrics between wake and sleep for each 24-hour recording. The first plot shows the difference between median levels of LF/HF and LFnu during wake and sleep, while the second plot also includes median HF. At a group level, there is a rough separation between the healthy controls, diabetics, and idiopathic gastroparesis patients. Across all three metrics, the healthy controls have the greatest magnitude of difference between sleep and wake, while the diabetics have the least, if any difference. Since all three differences were computed by subtracting the median levels during sleep from those during wake, the difference for both LF/HF and LFnu, which are more sympathetic metrics, is positive for healthy controls, while the difference in HF, which is parasympathetic, is negative. In all three cases, this suggests that in healthy controls, there is far more sympathetic activity during wake and greater parasympathetic activity during sleep, which agrees with known autonomic physiology [3]. In contrast, the diabetics seem to have little difference between wake and sleep in all three metrics, suggesting that appropriate circadian regulation of autonomic activity is impaired. The idiopathic gastroparesis patients have moderate difference between wake and sleep, suggesting less impairment than diabetics, but perhaps some dysregulation compared to healthy controls.

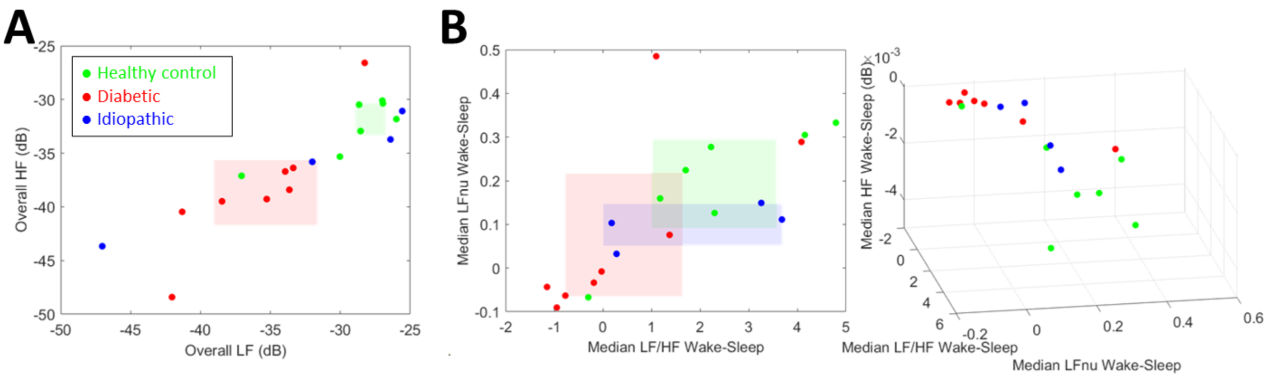


*Figure S1:* Summary of autonomic activity trends (A) Median LF vs median HF over 24 hours for all recordings (B) Comparison of difference in median LF/HF ratio between wake and sleep vs difference in median LFν between wake and sleep vs difference in median HF between wake and sleep for all recordings. All colored boxes show the 95% confidence interval of the mean in each dimension for that subgroup of subjects.

*Postprandial power*

The y-axis of Fig. S2A shows the area under the curve of normalized postprandial power around 0.05 Hz during the 4 hours after each isolated meal. Autonomic phenotypes are color-coded and specific GI phenotypes within them are denoted with different shapes. For example, diabetic non-gastroparetic meals are denoted with a diamond. Isolated meals with a gastric stimulator are denoted with squares. The gastric stimulators were not treated as a separate GI phenotype in later analyses in this study because of very few subjects, but there is a clear increase in postprandial power for isolated meals with gastric stimulators compared to without gastric stimulators for the same diagnosis, especially in the case of idiopathic gastroparesis patients. Overall, the 4-hr postprandial power is higher for healthy controls than for diabetic gastroparesis patients. Patients with diabetic non-gastroparetic diagnoses, such as dumping syndrome, also have increased postprandial power, suggesting an altered pathophysiology of disease.

*EGG Power in postprandial periods vs. fasting periods*

The x-axis of Fig. S2A shows the difference in area under the curve of normalized postprandial power around 0.05 Hz during the 4 hours after each isolated meal compared to the last 3 hours of overnight sleep in the same recording. The last 3 hours of overnight sleep were used as a proxy for a 'fasting' time period, since manual annotations of meals tend to be imprecise, and therefore relying on the annotated time period before a meal may be inaccurate and include some meal periods. In general, digestive activity should be at its peak in the postprandial period, especially compared to fasting (even accounting for migrating motor complexes [4]). From the x-axis of Fig. 4A, this is generally true for healthy controls, idiopathic patients with gastric stimulators, and diabetic non-gastroparesis patients. However, for most diabetic gastroparesis patients, even those with gastric stimulators, there is minimal difference between postprandial power and fasting power. This could be due to decreased intensity of postprandial power, which was the case in most situations, or inappropriately increased fasting activity, which was the case for one dumping syndrome subject.

The healthy control subject with the least separation between postprandial and fasting was a subject who was in their 80s, once again suggesting interesting hypothesis about the effects of aging on autonomic and digestive neurophysiology.

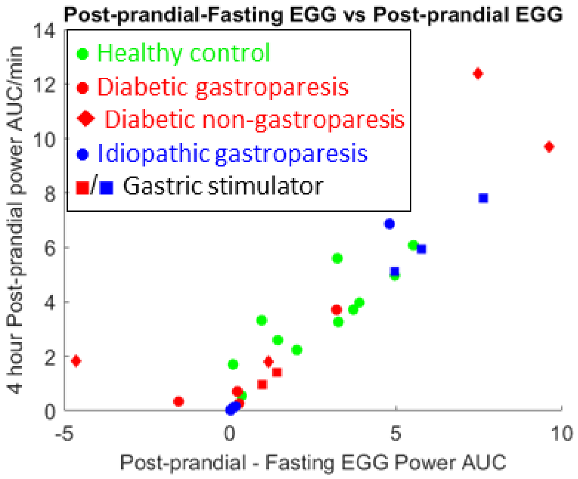


*Figure S2:* Summary of gastric motility activity patterns. Area under 4-hour postprandial normalized EGG power vs difference between area under postprandial and fasting (during sleep) EGG power for all isolated meals.

**References**


1. J. F. Kurniawan, B. Tjhia, V. M. Wu, A. Shin, N. L. J. Sit, T. Pham, A. Nguyen, C. Li, R. Kumar, M. Aguilar-Rivera, I. Lerman, D. C. Kunkel, T. P. Coleman (2021). “An Adhesive-Integrated Stretchable Ag/AgCl Electrode Array for Unobtrusive Monitoring of Gastric Neuromuscular Activity”, Advanced Materials Technologies. (Supp)
2. C. Brock, N. Jessen, B. Brock, P. E. Jakobsen, T. K. Hansen, J. M. Rantanen, S. Riahi, Y. K. Dimitrova, A. Dons-Jensen, Q. Aziz, A. M. Drewes, A. D. Farmer (2017). “Cardiac vagal tone, a non-invasive measure of parasympathetic tone, is a clinically relevant tool in Type 1 diabetes mellitus,” Diabet Med. 34(10):1428-1434. doi: 10.1111/dme.13421. Epub 2017 Aug 17. PMID: 28703868. (Supp)
3. R. Cabiddu, S. Cerutti, G. Viardot, S. Werner, A. M. Bianchi (2012). “Modulation of the Sympatho-Vagal Balance during Sleep: Frequency Domain Study of Heart Rate Variability and Respiration,” *Front Physiol.* **3**:45. doi: 10.3389/fphys.2012.00045. PMID: 22416233; PMCID: PMC3299415.
4. K. N. Browning, R. A. Travagli (2014). “Central nervous system control of gastrointestinal motility and secretion and modulation of gastrointestinal functions,” Comprehensive Physiology 4: 1339-68. (Supp)

**RAW DATA FIGS FOR ALL RECORDINGS**

For each recording, the HRV and gastric myoelectric indices are shown, with the spectrogram of vagal activity at the bottom. The x-axis of all the subplots, including the spectrogram, are aligned.

**Key**

**| Meals | Snacks | Going to sleep | Wake up | Bowel movement | Symptoms**

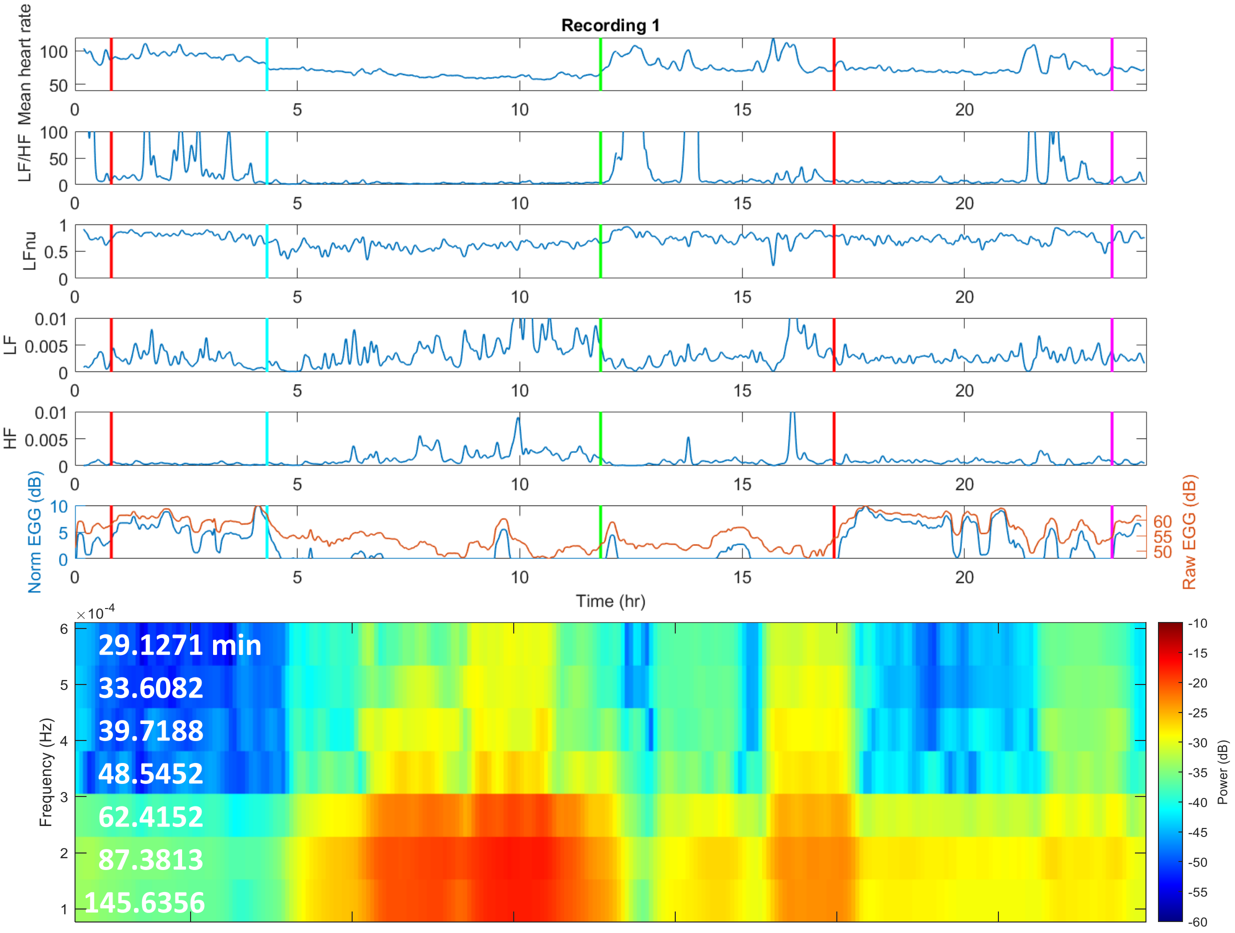


*Figure S3:* Autonomic (HRV) and gastric myoelectric information extracted from ECG and EGG data respectively, showing the mean heart rate, standard deviation of heart rate, sympathovagal balance, sympathetic and parasympathetic (vagal) modulation, normalized EGG power, and spectrogram of vagal activity at very low frequencies over the course of 24 hours for Recording 1

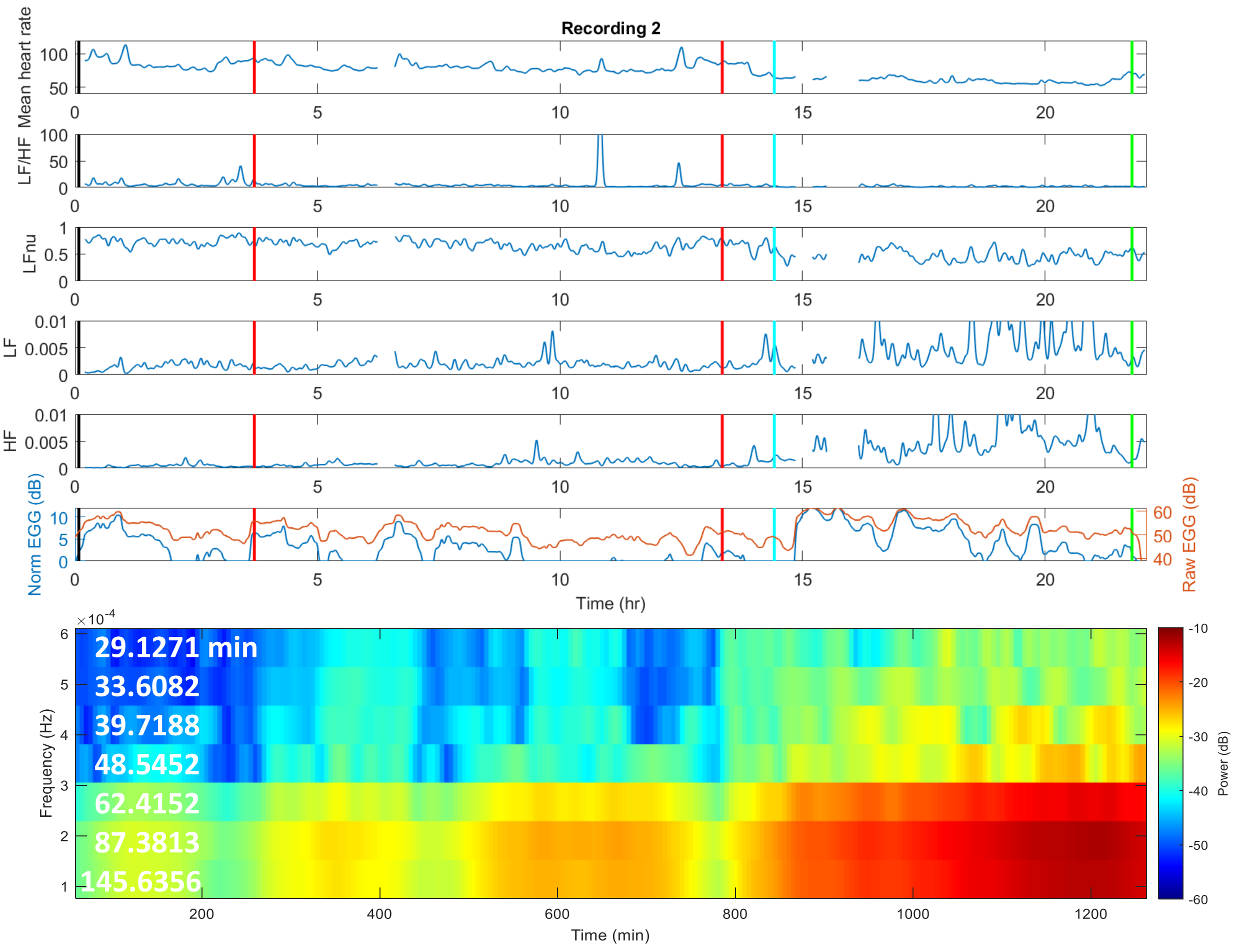


*Figure S4:* Autonomic (HRV) and gastric myoelectric information extracted from ECG and EGG data respectively, showing the mean heart rate, standard deviation of heart rate, sympathovagal balance, sympathetic and parasympathetic (vagal) modulation, normalized EGG power, and spectrogram of vagal activity at very low frequencies over the course of 24 hours for Recording 2

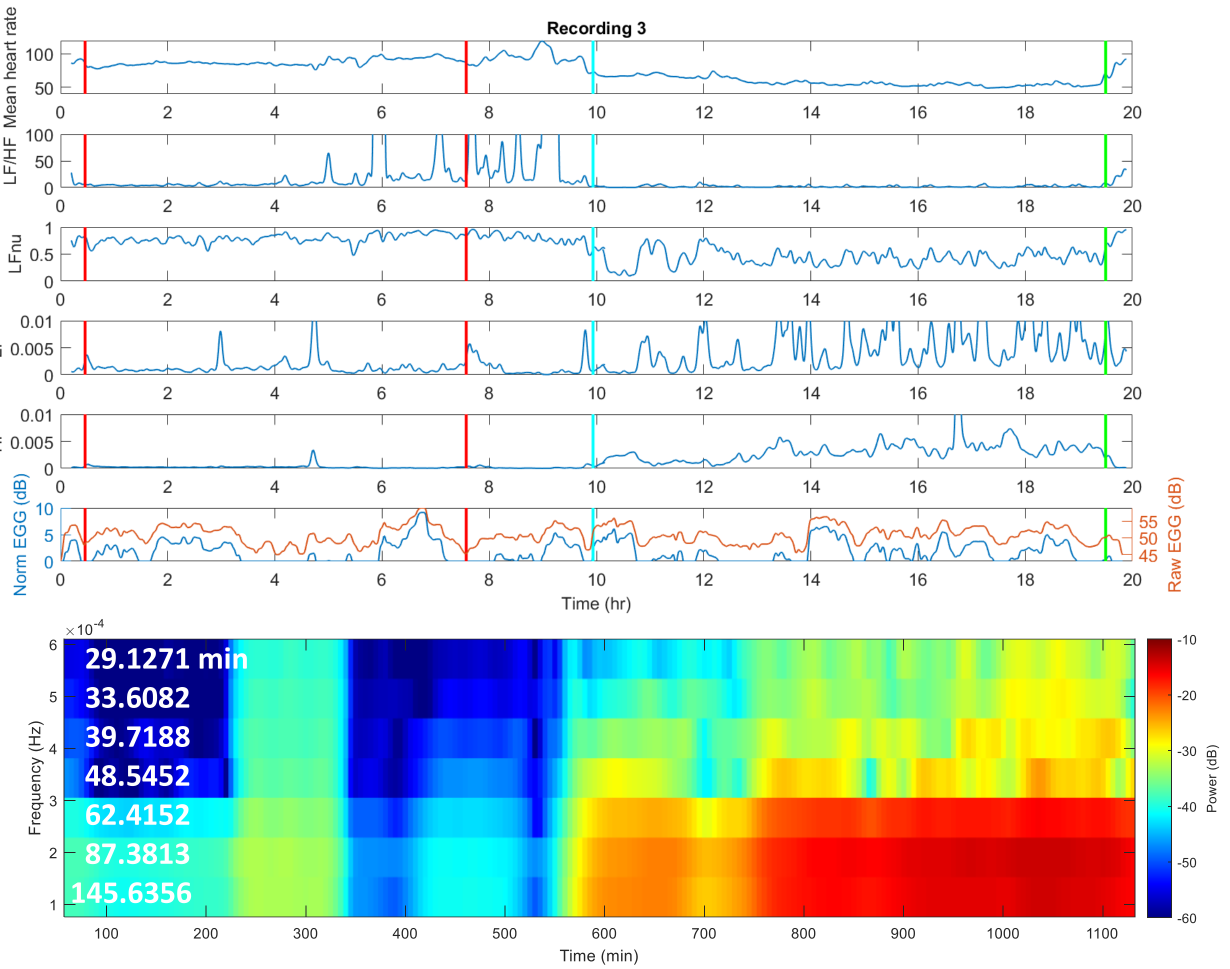


*Figure S5:* Autonomic (HRV) and gastric myoelectric information extracted from ECG and EGG data respectively, showing the mean heart rate, standard deviation of heart rate, sympathovagal balance, sympathetic and parasympathetic (vagal) modulation, normalized EGG power, and spectrogram of vagal activity at very low frequencies over the course of 24 hours for Recording 3

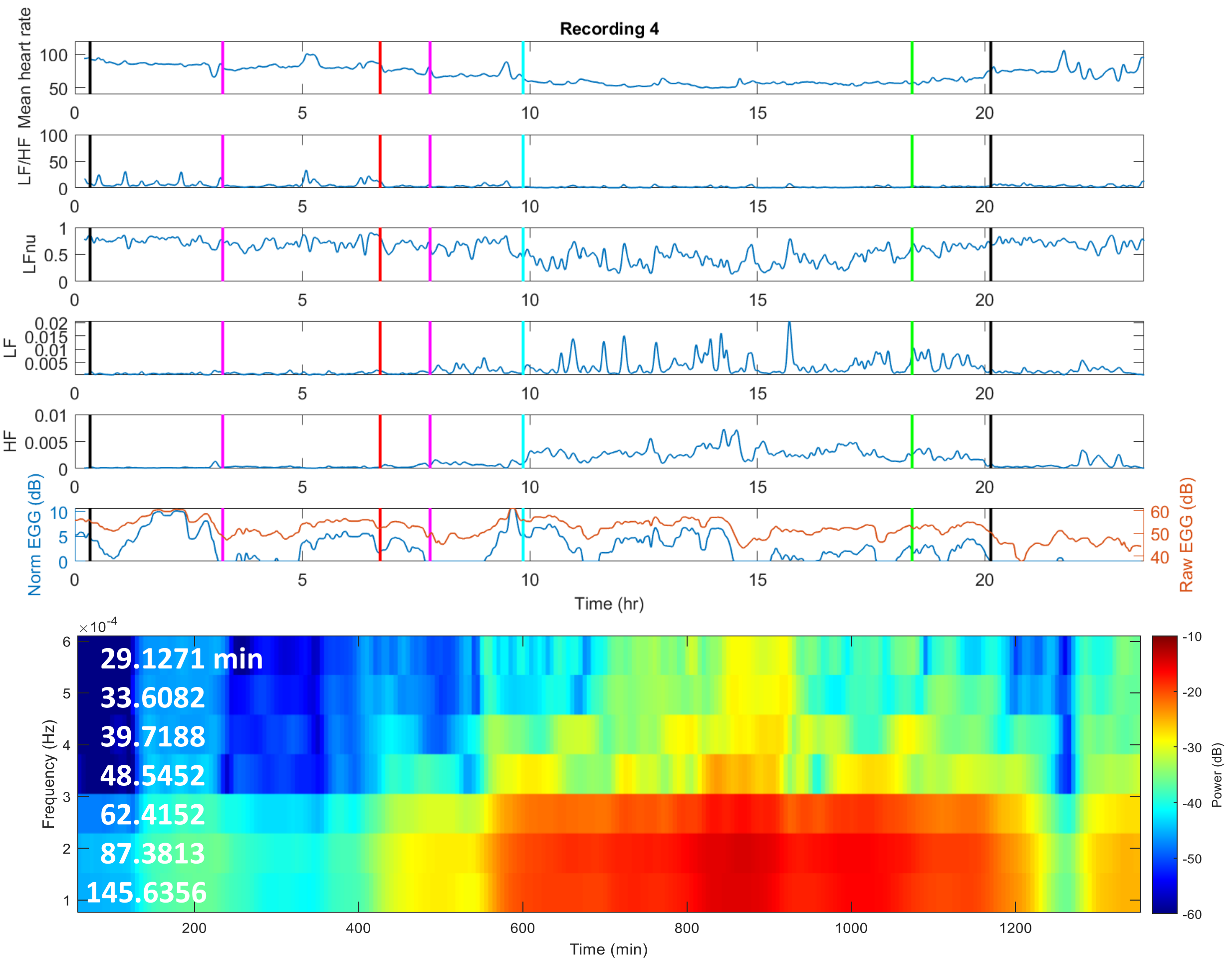


*Figure S6:* Autonomic (HRV) and gastric myoelectric information extracted from ECG and EGG data respectively, showing the mean heart rate, standard deviation of heart rate, sympathovagal balance, sympathetic and parasympathetic (vagal) modulation, normalized EGG power, and spectrogram of vagal activity at very low frequencies over the course of 24 hours for Recording 4

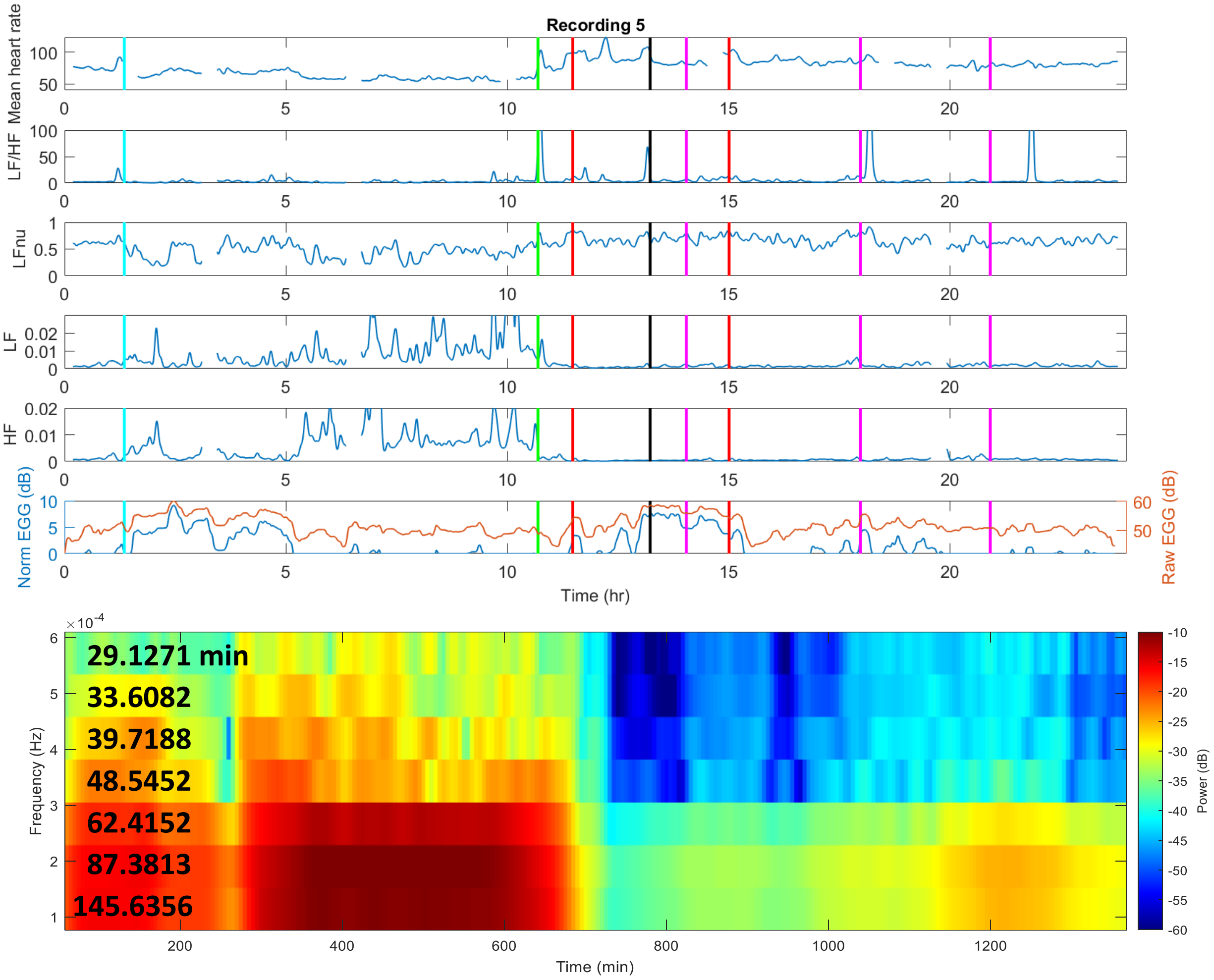


*Figure S7:* Autonomic (HRV) and gastric myoelectric information extracted from ECG and EGG data respectively, showing the mean heart rate, standard deviation of heart rate, sympathovagal balance, sympathetic and parasympathetic (vagal) modulation, normalized EGG power, and spectrogram of vagal activity at very low frequencies over the course of 24 hours for Recording 5

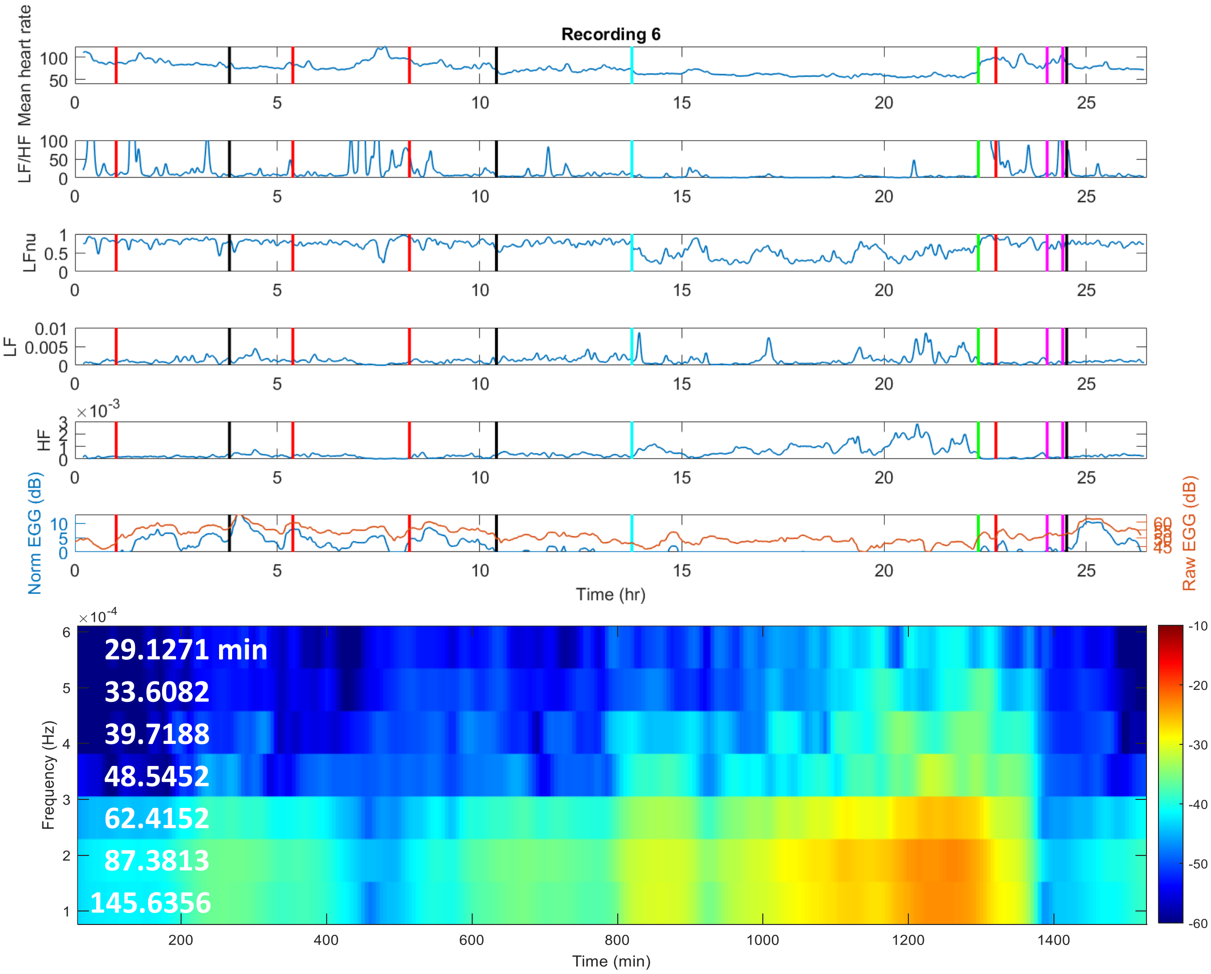


*Figure S8:* Autonomic (HRV) and gastric myoelectric information extracted from ECG and EGG data respectively, showing the mean heart rate, standard deviation of heart rate, sympathovagal balance, sympathetic and parasympathetic (vagal) modulation, normalized EGG power, and spectrogram of vagal activity at very low frequencies over the course of 24 hours for Recording 6

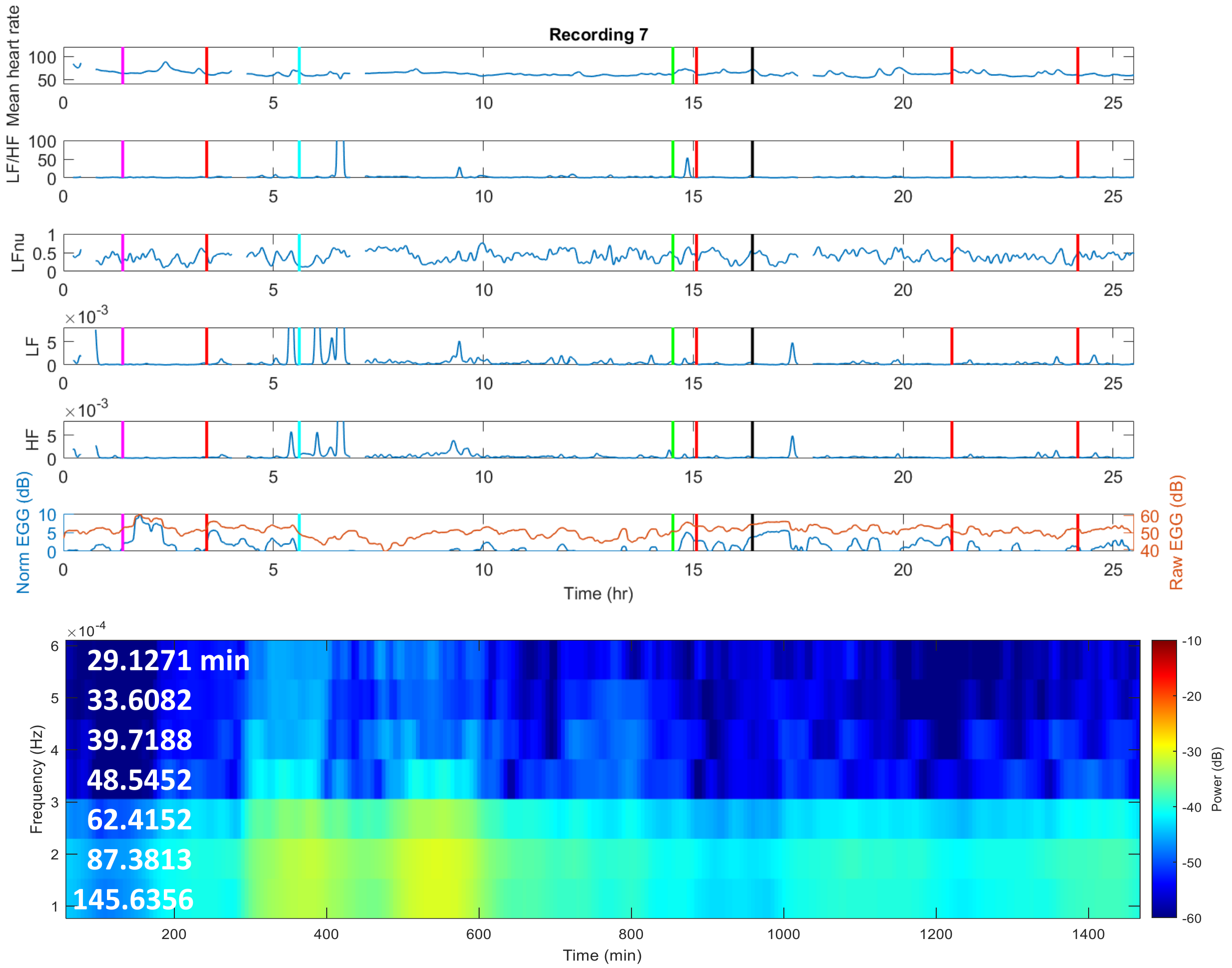


*Figure S9:* Autonomic (HRV) and gastric myoelectric information extracted from ECG and EGG data respectively, showing the mean heart rate, standard deviation of heart rate, sympathovagal balance, sympathetic and parasympathetic (vagal) modulation, normalized EGG power, and spectrogram of vagal activity at very low frequencies over the course of 24 hours for Recording 7

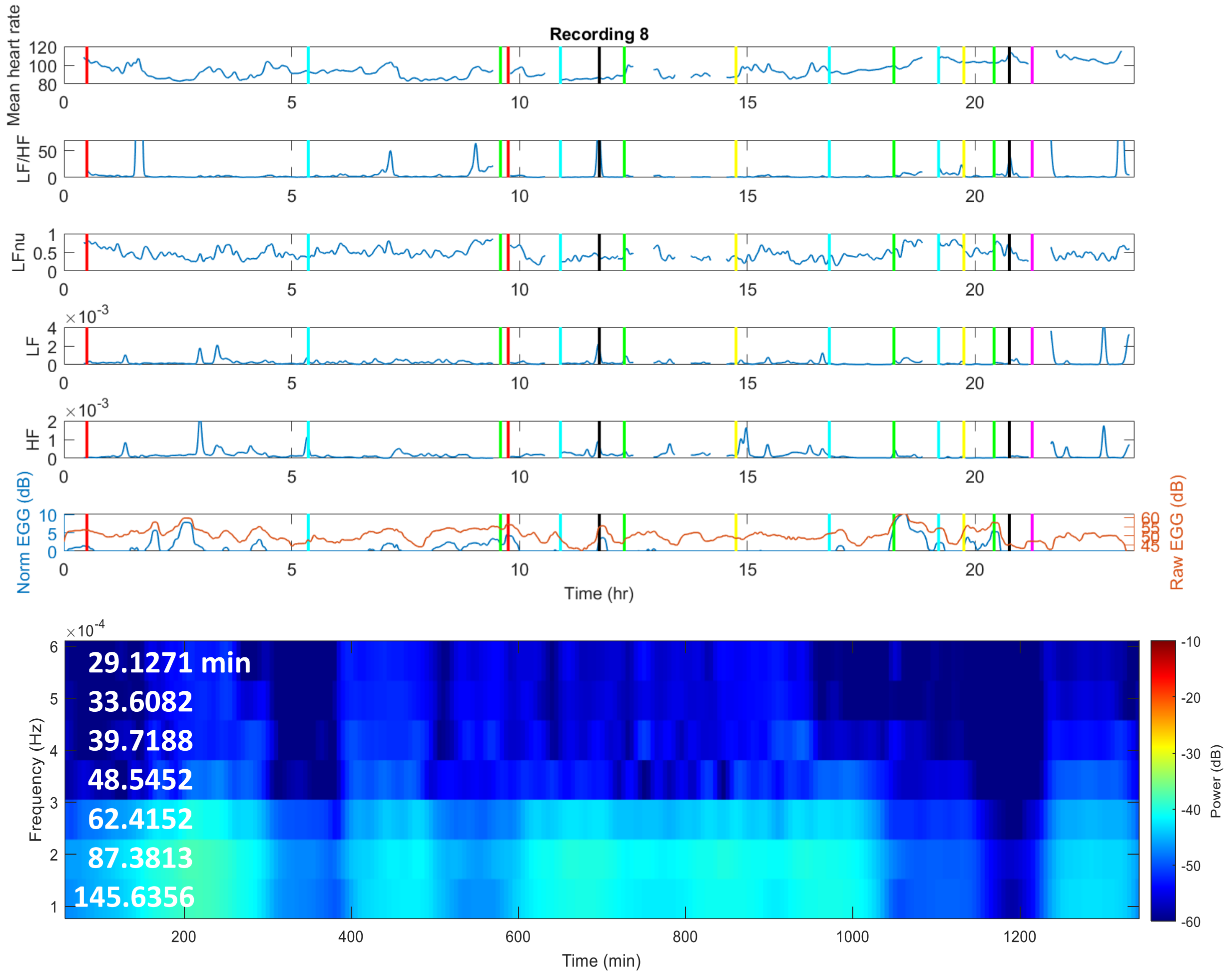


*Figure S10:* Autonomic (HRV) and gastric myoelectric information extracted from ECG and EGG data respectively, showing the mean heart rate, standard deviation of heart rate, sympathovagal balance, sympathetic and parasympathetic (vagal) modulation, normalized EGG power, and spectrogram of vagal activity at very low frequencies over the course of 24 hours for Recording 8

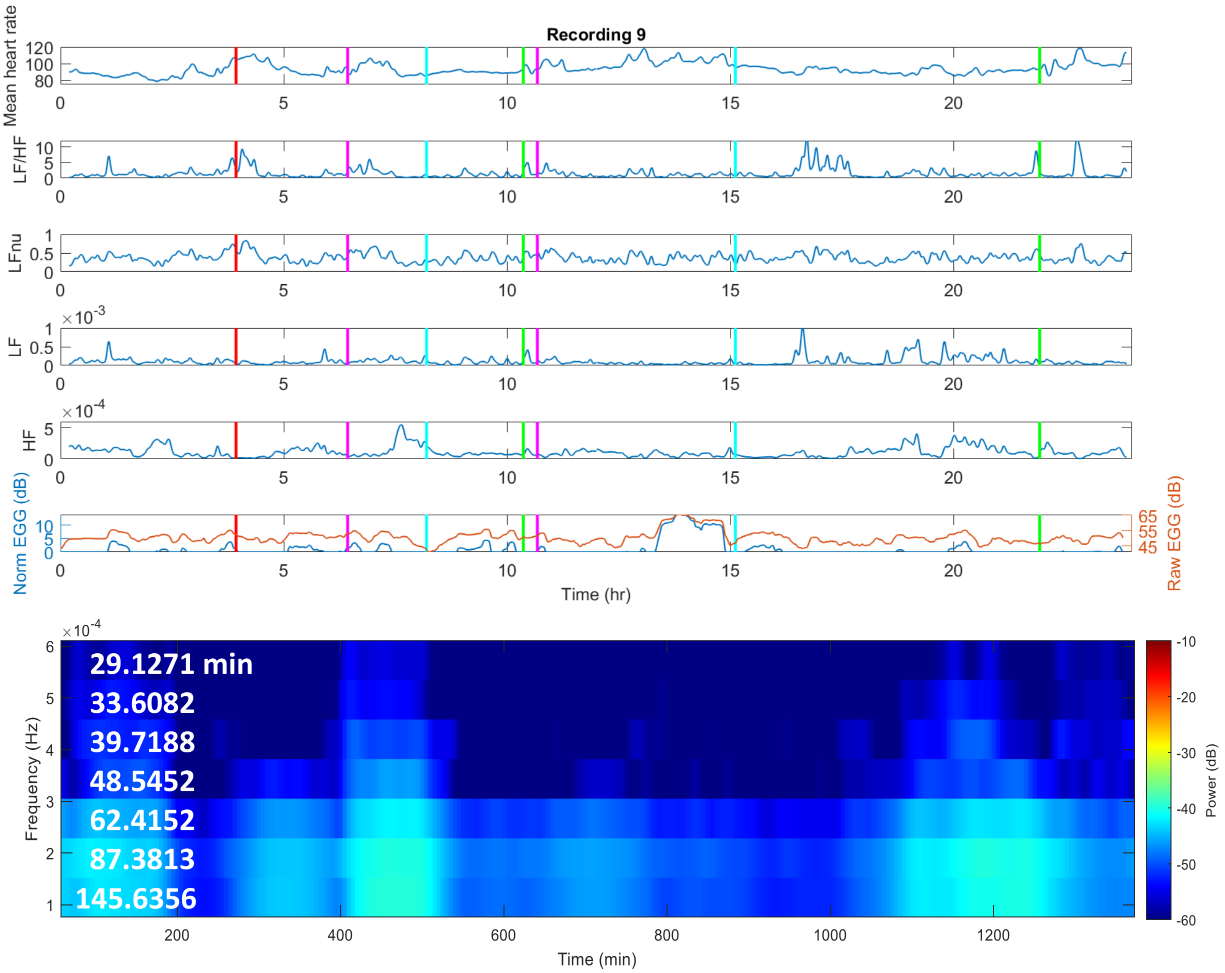


*Figure S11:* Autonomic (HRV) and gastric myoelectric information extracted from ECG and EGG data respectively, showing the mean heart rate, standard deviation of heart rate, sympathovagal balance, sympathetic and parasympathetic (vagal) modulation, normalized EGG power, and spectrogram of vagal activity at very low frequencies over the course of 24 hours for Recording 9

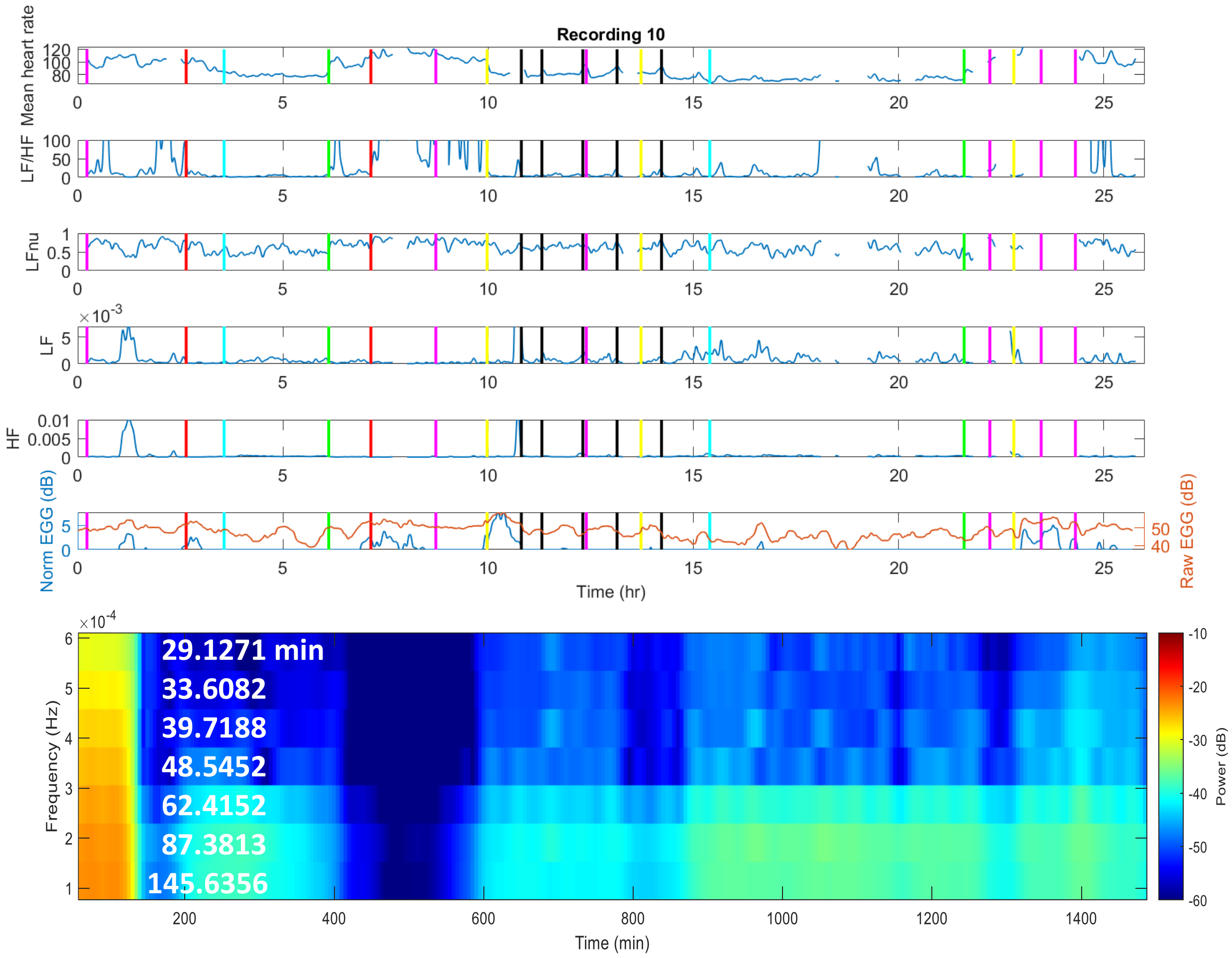


*Figure S12:* Autonomic (HRV) and gastric myoelectric information extracted from ECG and EGG data respectively, showing the mean heart rate, standard deviation of heart rate, sympathovagal balance, sympathetic and parasympathetic (vagal) modulation, normalized EGG power, and spectrogram of vagal activity at very low frequencies over the course of 24 hours for Recording 10

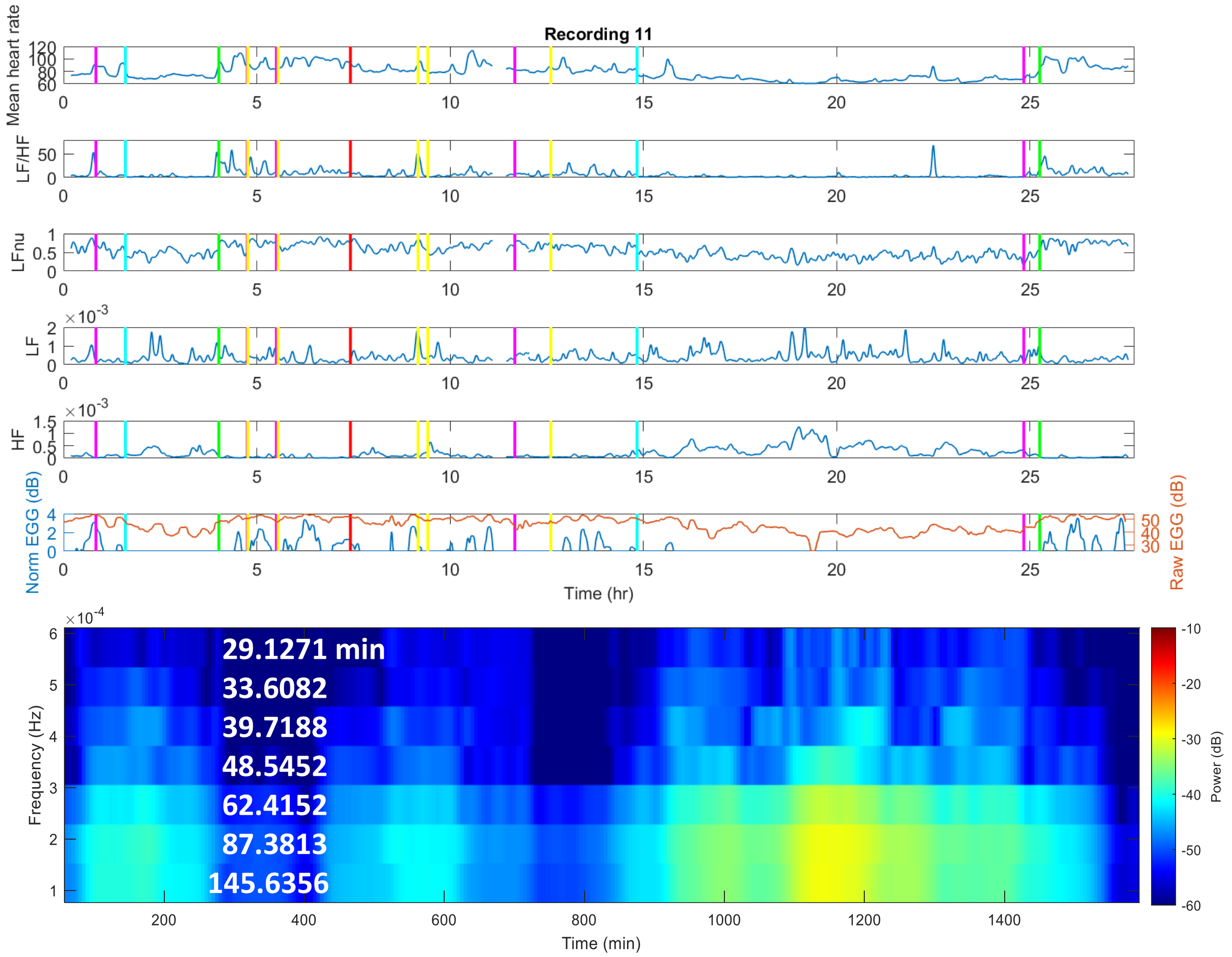


*Figure S13:* Autonomic (HRV) and gastric myoelectric information extracted from ECG and EGG data respectively, showing the mean heart rate, standard deviation of heart rate, sympathovagal balance, sympathetic and parasympathetic (vagal) modulation, normalized EGG power, and spectrogram of vagal activity at very low frequencies over the course of 24 hours for Recording 11

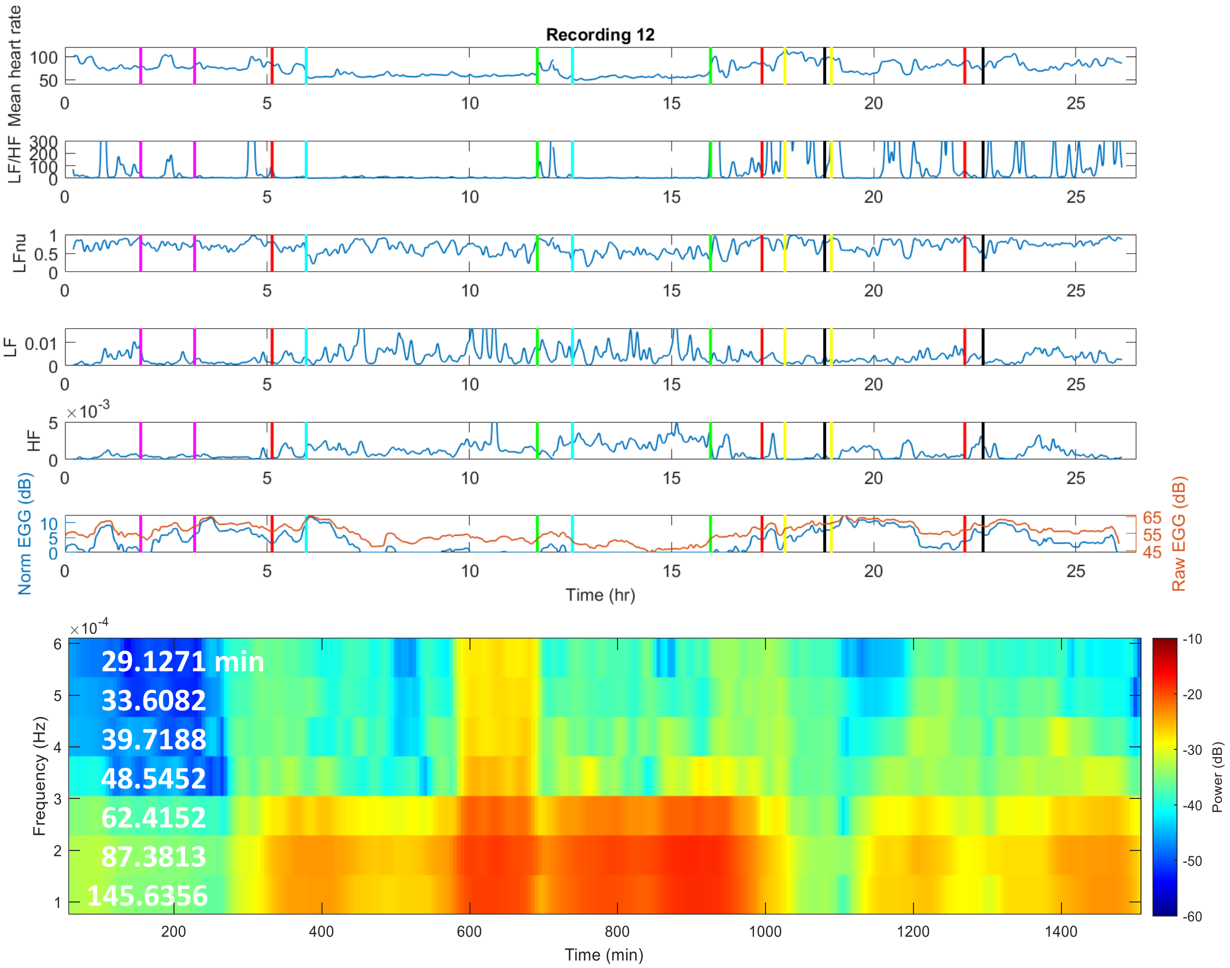


*Figure S14:* Autonomic (HRV) and gastric myoelectric information extracted from ECG and EGG data respectively, showing the mean heart rate, standard deviation of heart rate, sympathovagal balance, sympathetic and parasympathetic (vagal) modulation, normalized EGG power, and spectrogram of vagal activity at very low frequencies over the course of 24 hours for Recording 12

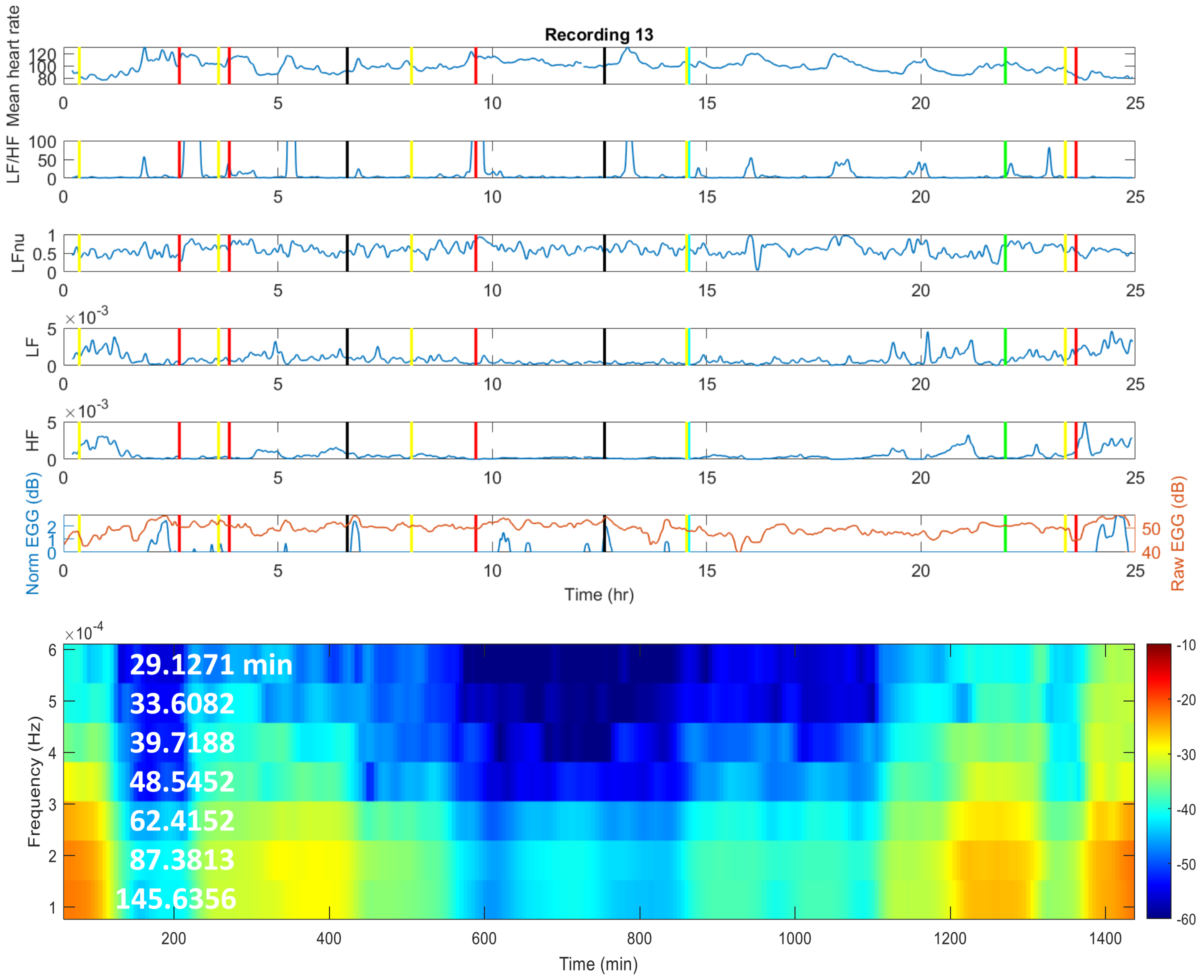


*Figure S15:* Autonomic (HRV) and gastric myoelectric information extracted from ECG and EGG data respectively, showing the mean heart rate, standard deviation of heart rate, sympathovagal balance, sympathetic and parasympathetic (vagal) modulation, normalized EGG power, and spectrogram of vagal activity at very low frequencies over the course of 24 hours for Recording 13

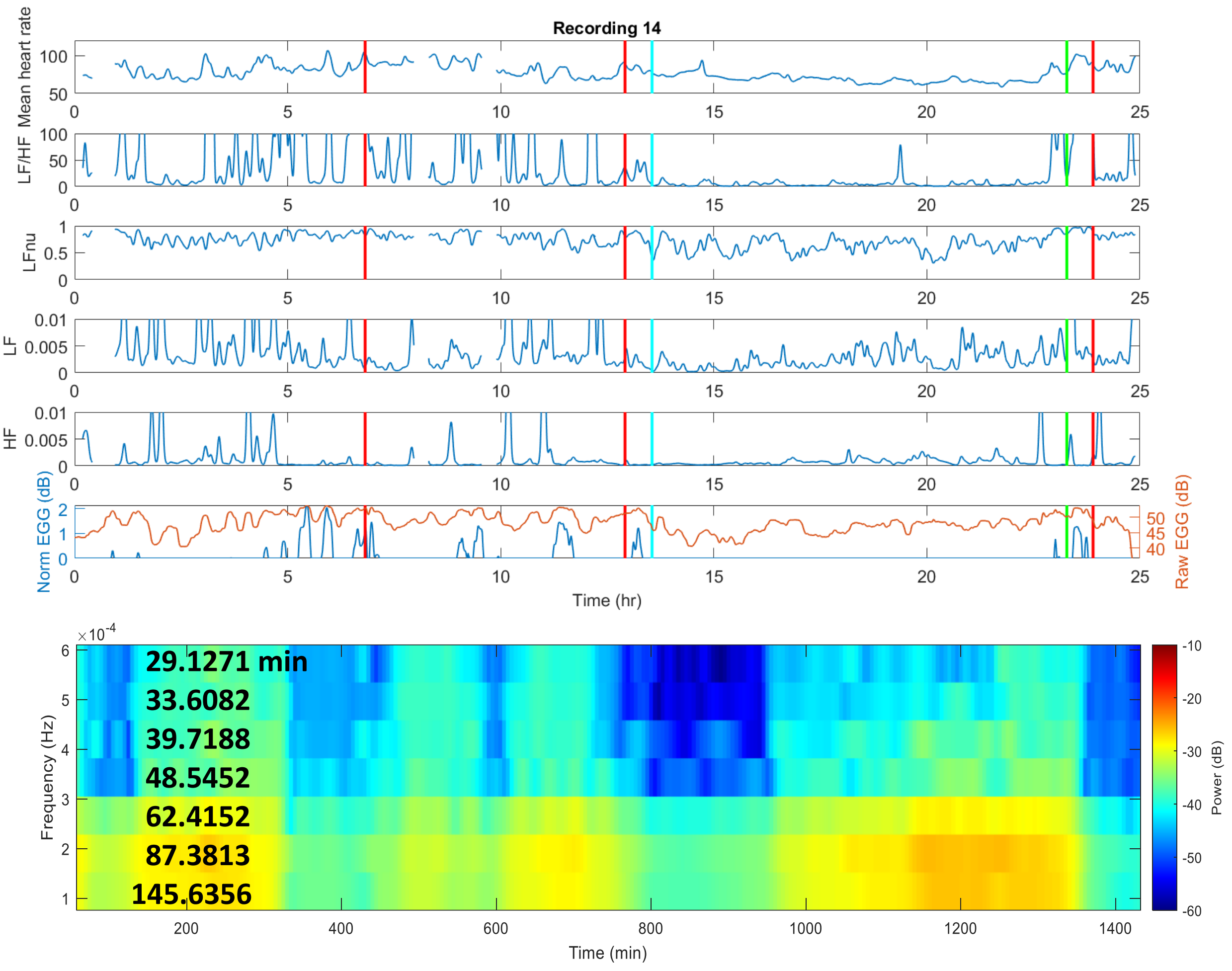


*Figure S16:* Autonomic (HRV) and gastric myoelectric information extracted from ECG and EGG data respectively, showing the mean heart rate, standard deviation of heart rate, sympathovagal balance, sympathetic and parasympathetic (vagal) modulation, normalized EGG power, and spectrogram of vagal activity at very low frequencies over the course of 24 hours for Recording 14

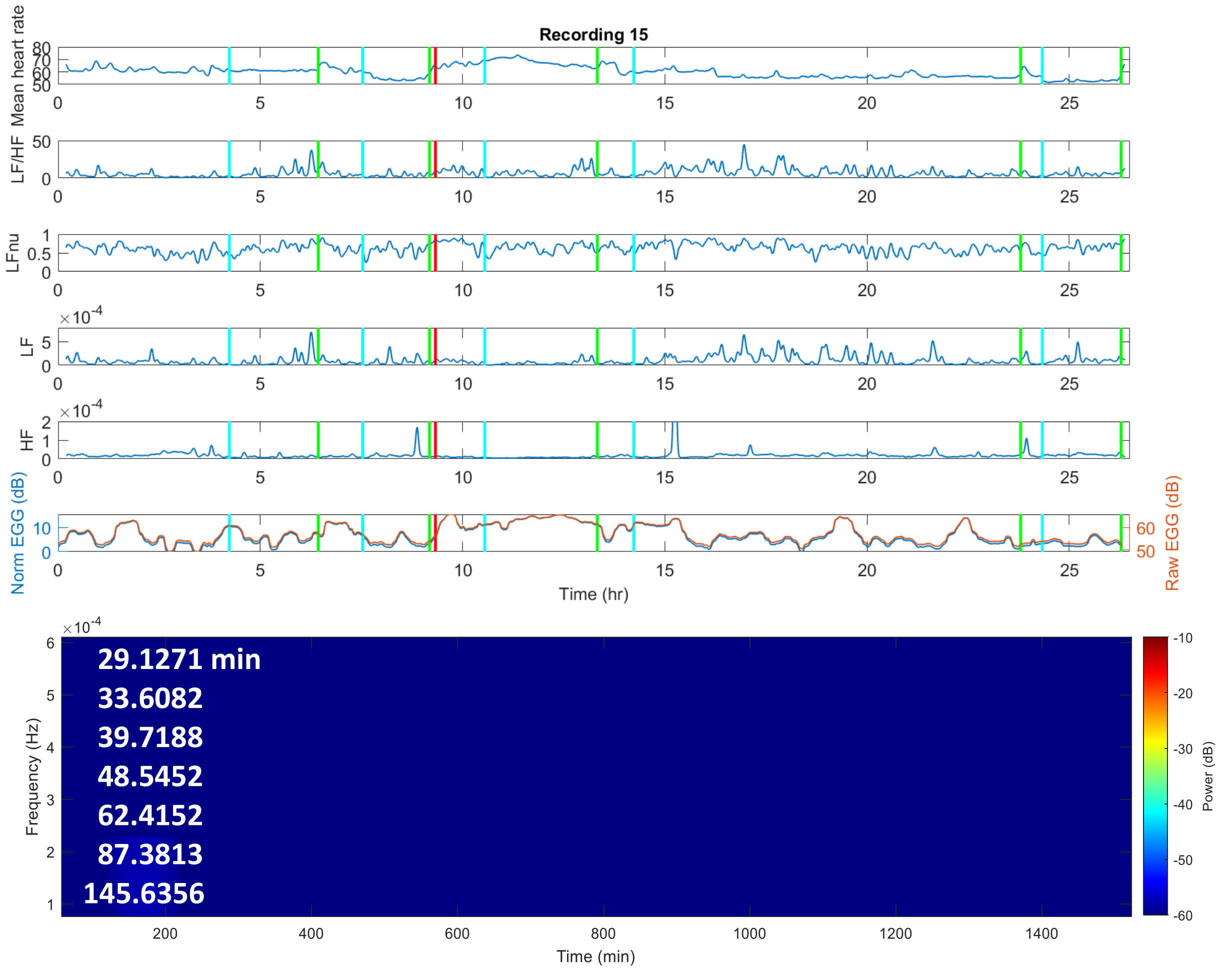


*Figure S17:* Autonomic (HRV) and gastric myoelectric information extracted from ECG and EGG data respectively, showing the mean heart rate, standard deviation of heart rate, sympathovagal balance, sympathetic and parasympathetic (vagal) modulation, normalized EGG power, and spectrogram of vagal activity at very low frequencies over the course of 24 hours for Recording 15

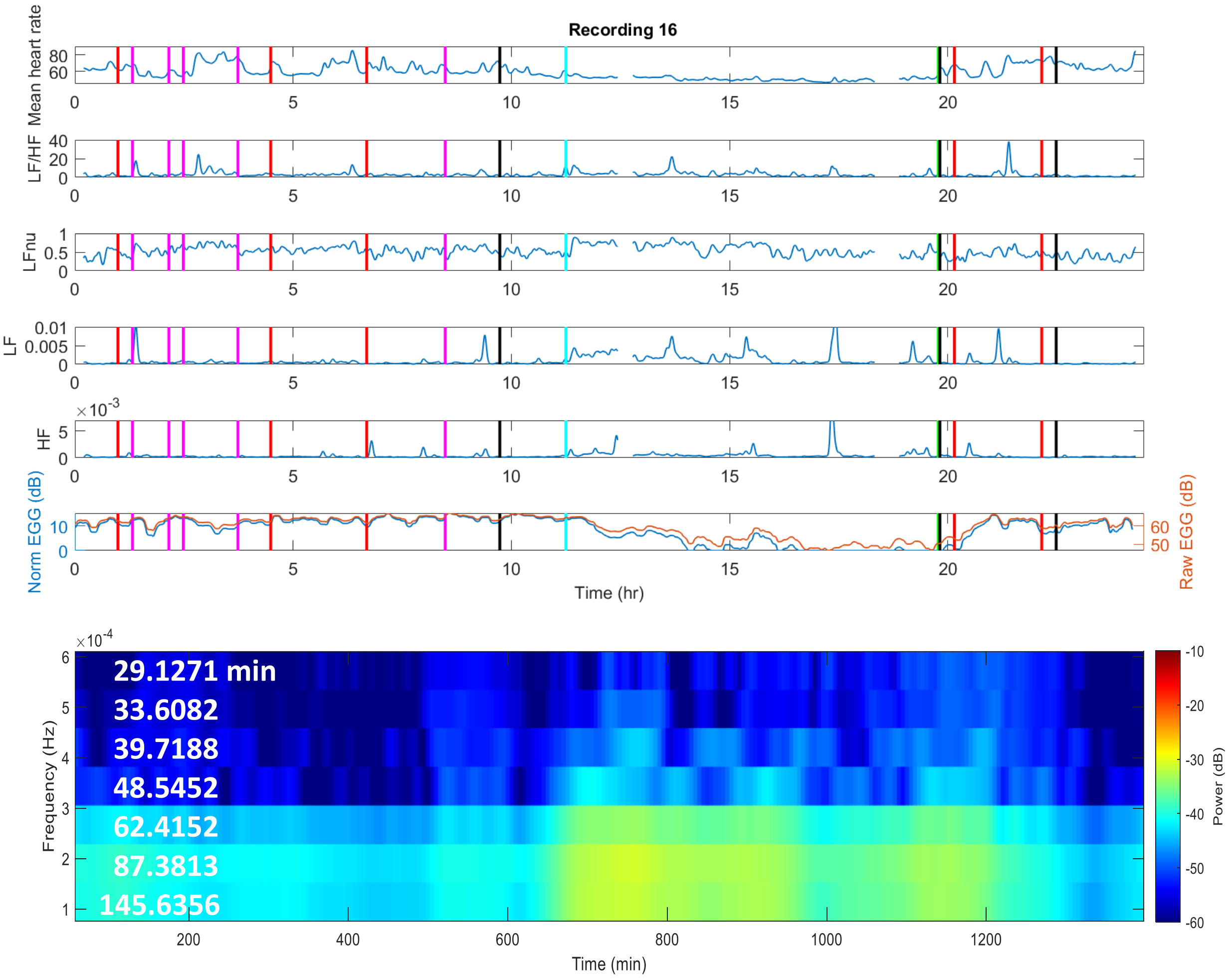


*Figure S18:* Autonomic (HRV) and gastric myoelectric information extracted from ECG and EGG data respectively, showing the mean heart rate, standard deviation of heart rate, sympathovagal balance, sympathetic and parasympathetic (vagal) modulation, normalized EGG power, and spectrogram of vagal activity at very low frequencies over the course of 24 hours for Recording 16

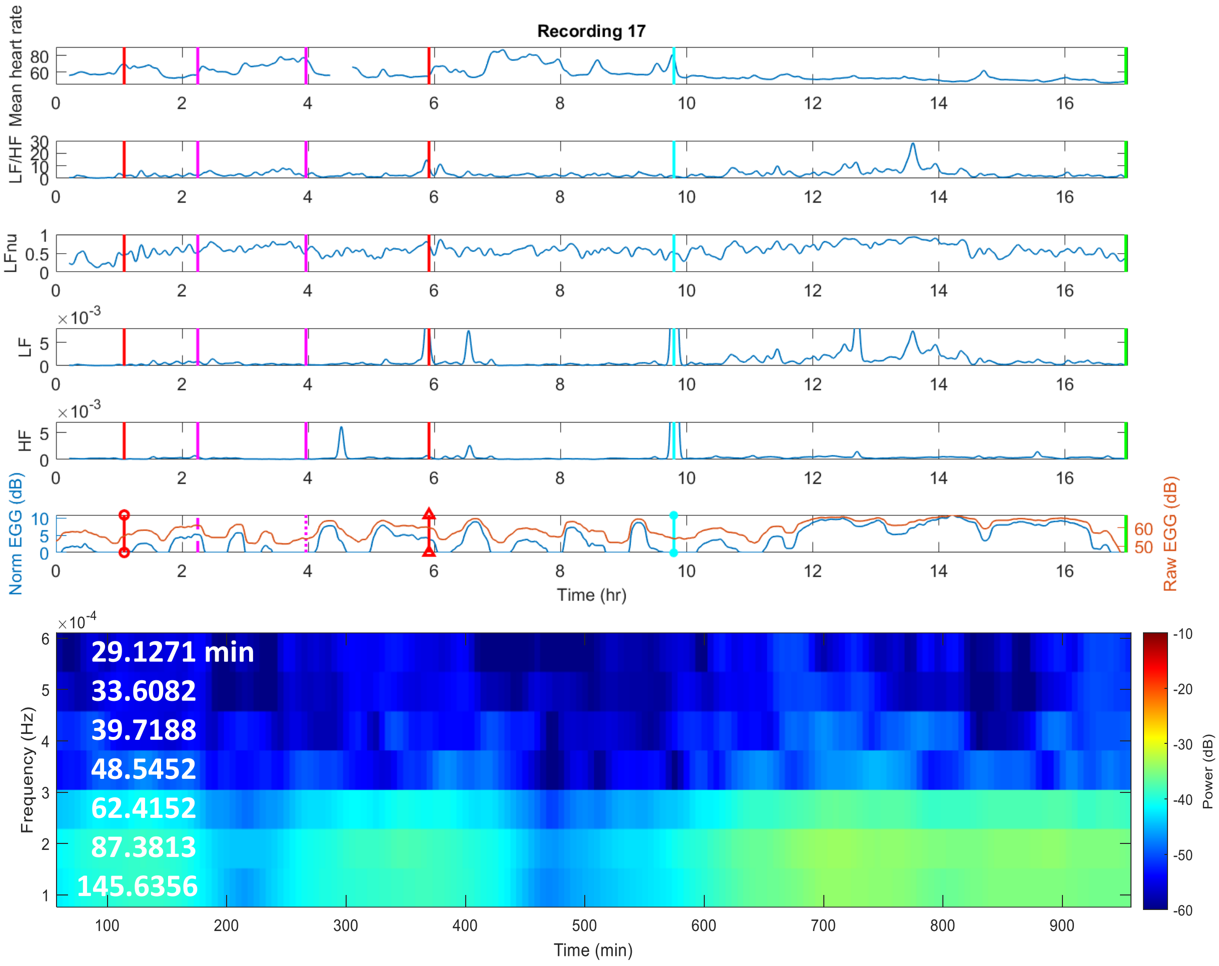


*Figure S19:* Autonomic (HRV) and gastric myoelectric information extracted from ECG and EGG data respectively, showing the mean heart rate, standard deviation of heart rate, sympathovagal balance, sympathetic and parasympathetic (vagal) modulation, normalized EGG power, and spectrogram of vagal activity at very low frequencies over the course of 24 hours for Recording 17

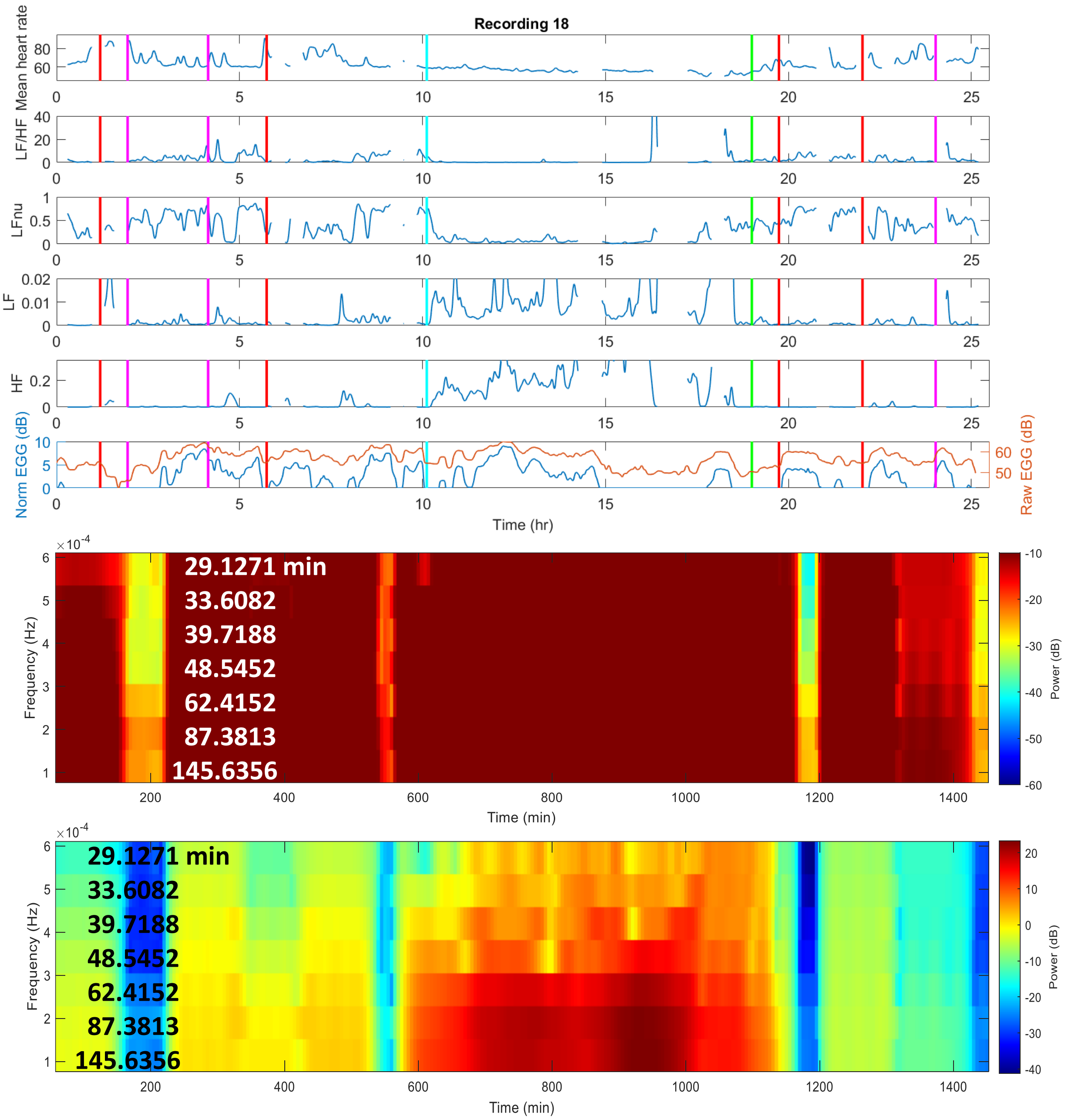


*Figure S20:* Autonomic (HRV) and gastric myoelectric information extracted from ECG and EGG data respectively, showing the mean heart rate, standard deviation of heart rate, sympathovagal balance, sympathetic and parasympathetic (vagal) modulation, normalized EGG power, and spectrogram of vagal activity at very low frequencies over the course of 24 hours for Recording 18

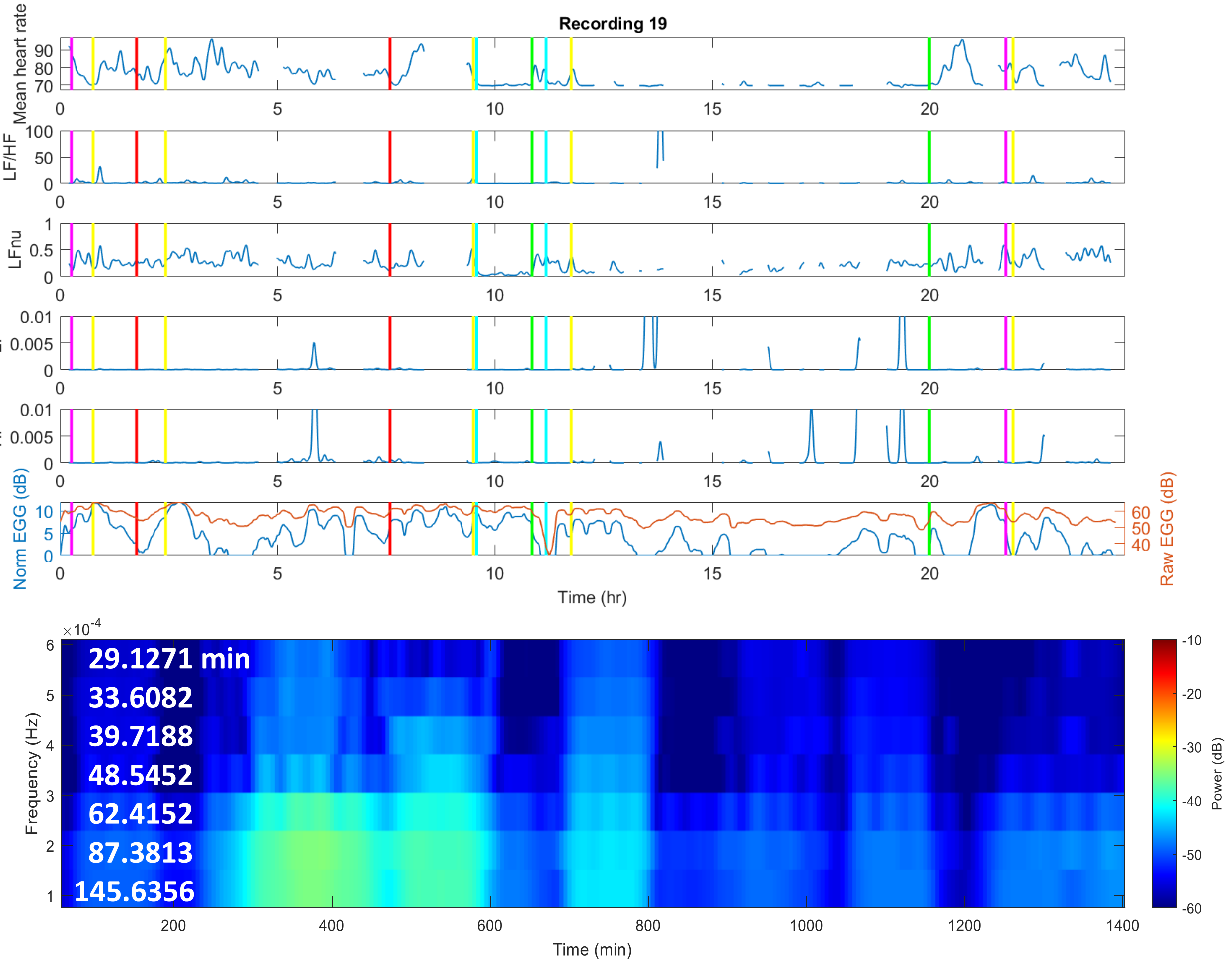


*Figure S21:* Autonomic (HRV) and gastric myoelectric information extracted from ECG and EGG data respectively, showing the mean heart rate, standard deviation of heart rate, sympathovagal balance, sympathetic and parasympathetic (vagal) modulation, normalized EGG power, and spectrogram of vagal activity at very low frequencies over the course of 24 hours for Recording 19

**BOOTSTRAPPED CONFIDENCE INTERVALS**

**Fig S1A**

| Recording | 95% CI for Median Overall LF | 95% CI for Median Overall HF |
|---|---|---|
| **1** | -25.9941 -25.9824 | -31.8400 -31.8147 |
| **2** | -26.9984 -26.9827 | -30.1189 -30.0764 |
| **3** | -28.5583 -28.5371 | -32.9901 -32.8943 |
| **4** | -28.6618 -28.6440 | -30.5247 -30.4188 |
| **5** | -26.9568 -26.9364 | -30.3730 -30.3448 |
| **6** | -30.0365 -30.0226 | -35.3338 -35.3150 |
| **7** | -37.0740 -37.0518 | -37.1159 -37.0979 |
| **8** | -38.4713 -38.4493 | -39.5011 -39.4726 |
| **9** | -41.3229 -41.3022 | -40.4676 -40.4522 |
| **10** | -33.6568 -33.6280 | -38.4231 -38.3947 |
| **11** | -35.2713 -35.2567 | -39.2843 -39.2677 |
| **12** | -25.5756 -25.5573 | -31.0896 -31.0614 |
| **13** | -32.0147 -31.9898 | -35.8145 -35.7894 |
| **14** | -26.4163 -26.3982 | -33.7298 -33.7096 |
| **15** | -42.0527 -42.0270 | -48.4170 -48.4040 |
| **16** | -33.9522 -33.9275 | -36.7161 -36.6977 |
| **17** | -33.3785 -33.3561 | -36.3789 -36.3615 |
| **18** | -28.2883 -28.2194 | -26.6229 -26.5322 |
| **19** | -47.0483 -47.0043 | -43.6872 -43.6415 |

**Fig S1B**

| Recording | 95% CI for Difference in Median LF/HF Wake-Sleep | 95% CI for Difference in Median LFnu Wake-Sleep | 95% CI for Difference in Median HF Wake-Sleep |
|---|---|---|---|
| **1** | 2.2868 2.3086 | 0.1254 0.1266 | -7.6875e-4 -7.5478e-4 |
| **2** | 1.6984 1.7109 | 0.2234 0.2252 | -0.0027 -0.0027 |
| **3** | 4.7796 4.8031 | 0.3318 0.3339 | -0.0027 -0.0027 |
| **4** | 2.2176 2.2334 | 0.2759 0.2785 | -0.0025 -0.0025 |
| **5** | 1.1725 1.1854 | 0.1585 0.1605 | -0.0048 -0.0047 |
| **6** | 4.1408 4.1719 | 0.3025 0.3074 | -5.9901e-4 -5.9441e-4 |
| **7** | -0.3038 -0.2924 | -0.0683 -0.0659 | -1.4761e-4 -1.4517e-4 |
| **8** | -0.1914 -0.1779 | -0.0348 -0.0324 | 6.0771e-5 6.2042e-5 |
| **9** | -0.0307 -0.0216 | -0.0093 -0.0065 | 5.5175e-6 6.0248e-6 |
| **10** | 1.3540 1.3891 | 0.0747 0.0768 | -1.5525e-4 -1.5408e-4 |
| **11** | 4.0714 4.0977 | 0.2880 0.2897 | -2.5299e-4 -2.5050e-4 |
| **12** | 3.2355 3.2840 | 0.1482 0.1502 | -0.0012 -0.0012 |
| **13** | 0.2759 0.2904 | 0.0318 0.0335 | -1.7446e-5 -1.3382e-5 |
| **14** | 3.6593 3.7072 | 0.1105 0.1119 | -1.5182e-4 -1.4878e-4 |
| **15** | -1.1658 -1.1306 | -0.0440 -0.0426 | -9.6651e-8 -1.0468e-8 |
| **16** | -0.9582 -0.9306 | -0.0914 -0.0892 | -2.3814e-4 -2.3572e-4 |
| **17** | -0.7829 -0.7601 | -0.0640 -0.0623 | -2.0902e-4 -2.0724e-4 |
| **18** | 1.0924 1.1049 | 0.4838 0.4866 | -0.1484 -0.1473 |
| **19** | 0.1804 0.1832 | 0.1024 0.1041 | 3.3167e-05 3.4219e-05 |
